# HIV Transmission in a Declining African Epidemic

**DOI:** 10.64898/2026.04.29.26350859

**Authors:** Griffin J Bell, M Kate Grabowski, Josephine Mpagazi, Francesco Di Lauro, Aleya Khalifa, Anthony Ndyanabo, Hadijja Nakawooya, Joseph Kagaayi, Godfrey Kigozi, Gertrude Nakigozi, Ronald M Galiwango, Grace Kigozi, Michael A Martin, Luca Ferretti, Christophe Fraser, David Bonsall, Lucie Abeler-Dörner, Tanya Golubchik, Aaron AR Tobian, Laura K. Beres, Caitlin Kennedy, Justin Lessler, Thomas C Quinn, Steven J Reynolds, Maria J Wawer, Ronald H Gray, David Serwadda, Larry W Chang, Robert Ssekubugu

## Abstract

**Background:** Novel HIV prevention interventions such as long-acting pre-exposure prophylaxis (PrEP) could substantially reduce HIV transmission in Africa. However, efficient implementation in high-prevalence settings where incidence has declined requires an understanding of the contemporary dynamics driving new infections.

**Methods:** We identified incident HIV cases from a longitudinal, population-based cohort in Uganda. We individually matched cases to HIV-negative controls; traced and enrolled reported sexual partners; and enrolled female sex workers (FSWs) from reported venues. Conditional logistic regression, transmission modeling, and phylogenetics were used to characterize transmission networks.

**Findings:** From 2021-2024, 38,899 HIV tests among 22,255 people identified 187 people with incident infections (47.6% male); 164 (88%) were enrolled and matched to 164 HIV-negative controls. Overall, 593 non-sex-worker partners (371 enrolled,62.6%), 146 FSW partners (21 enrolled,14.4%), and 28 venues (208 FSWs enrolled) were reported. Incident infection was most strongly predicted by partnership with a FSW (odds ratio:15.5; 95%CI:3.7-64.8), identified in 43.0% of male cases versus 6.3% of controls. Men with FSW partners had larger sexual networks than men without (median:6 vs 2 partners), and 91.2% of men with FSW partners also had non-sex-worker partners. Transmission modeling attributed 34.4% (95%CI:31.5-36.8%) of all male infections and 80.0% (95%CI:73.2-84.4%) of infections among male clients to sex with FSWs. Oral PrEP use among HIV-negative partners of incident cases was low (8.9% in women; 2.1% in men).

**Interpretation:** Men with FSW partners accounted for a substantial share of incident HIV infections and had markedly higher odds of infection than men without such partnerships. Together with the high potential for onward transmission within male client networks, these findings suggest that inclusion of male clients in long-acting HIV prevention strategies could be highly efficient and impactful.

**Funding:** National Institutes of Health, United States; Gates Foundation; National Health and Medical Research Council, Australia

## INTRODUCTION

Since 2010, HIV incidence in eastern and southern Africa (ESA) has declined by nearly 60% (1). In 2024, 93% of people living with HIV in ESA were diagnosed, 91% of people diagnosed were on treatment, and 95% of people on treatment were virally suppressed (1). Despite these achievements, there were 490,000 new HIV infections, accounting for 37% of global incident infections (1). In the context of declining incidence (2) and constrained international funding for HIV treatment and prevention (3), effectively reaching sexual networks where transmission persists is critical for achieving HIV epidemic control.

However, identifying these networks remains challenging in historically generalized epidemics that are now characterized by low incidence. Some predict that as epidemics decline, incidence will increasingly cluster in key populations at elevated HIV risk (e.g., female sex workers) (4,5). However, data from across ESA suggest that a minority of new infections occur in such high-risk populations (1,6,7), with no notable shifts over the past decade (6,8). Moreover, phylogenetic studies depict a diffuse epidemic with few large transmission clusters (9–11), and conflict as to whether high-HIV-prevalence groups and geographic regions are sources or sinks of transmission (12–14). Taken together, these findings suggest that transmission may remain broadly distributed across populations.

Uncertainty about the structure of ongoing transmission in declining, low-incidence epidemics partly reflects limitations of prior study designs. Most prior studies were conducted before widespread ART scale-up and incidence declines, and rarely integrated key populations with the broader community. Moreover, population-based cohorts often underrepresent individuals most likely to sustain transmission because they are harder to reach and less engaged in care (15–17). Although assisted partner notification (APN) programs aim to capture transmission networks of newly diagnosed people (18), they typically lack well-matched controls and are subject to selection bias, limiting inference about network-level risk factors and population-level transmission dynamics. As a result, the extent to which transmission remains generalized versus concentrated within specific networks remains unclear.

Here, we conducted a matched, case-control and contact-tracing study nested within a longitudinal, population-representative cohort in southern Uganda: a setting with high levels of viral suppression and declining HIV incidence (11,15,19), representative of many later-stage African epidemics (2). We enrolled sexual partners of population-representative incident HIV cases and HIV-negative controls, and characterized the prevalence of HIV, viremia, sex work, and intervention uptake across these networks to inform efficient implementation of interventions, like long-acting prevention.

## METHODS

### Parent Study Description

This matched case-control study was nested within rounds 20 (January 2021-March 2023) and 21 (April 2023-June 2025) of the Rakai Community Cohort Study (RCCS), a population-based cohort of all persons aged ≥15 years in 34 communities in southern Uganda, described in the Supplemental Methods and elsewhere (15,20). All participants were provided HIV services according to national guidelines and linked to follow-up care with the regional HIV control program (Supplemental Methods).

### RCCS Laboratory Methods

HIV serostatus was assessed using a validated rapid testing algorithm with confirmation by ELISA (21). Viral load testing was performed using the Abbott M2000rt (Abbott Laboratories, Abbott Park, IL) and HIV serologic recency testing was done using the Asante HIV-1 Rapid Recency assay (Sedia Biosciences Corporation, Portland, OR). Participants with HIV were considered virally suppressed with <200 copies per mL and viremic with ≥200 copies.

### Incident HIV Cases and Matched Controls

New HIV diagnoses identified between January 2021 and February 2024 in the RCCS were eligible for this study and were classified as incident by satisfying any of the following: 1) seroconversion documented by longitudinal serologic HIV testing (an HIV seropositive test following a negative test in the prior two RCCS rounds); 2) high level viremia (>1000 copies/ml) with a positive cross-sectional serologic recency testing result (22); and 3) high level viremia with a self-reported last-negative HIV test result in the past year. HIV-negative controls, enrolled from the RCCS, were matched to cases based on sex (male or female), age (within 2 years), community type (Lake Victoria fishing or inland) of residence, and religion (Christian or other).

### Partner and Venue Reporting

Sexual partners in the previous 24 months were reported by cases and controls, and enrollment was attempted for all partners with sufficient contact information. Many FSW partners were untraceable, as participants often reported venue-based encounters without identifying details. In contrast to many assisted partner notification studies, which report tracing success among partners with contact information, our estimates include all reported partners. The study protocol was modified in May 2022 to include reporting of FSW venues in response to frequent reporting of untraceable FSW partners. Managers at reported venues provided a list of FSWs present on the same dates as the index participant. From that list, consenting FSWs were enrolled. All enrolled sexual partners and venue-based FSWs were counseled and tested for HIV and referred for appropriate HIV services, and venues were reported to the regional HIV program.

### Survey-Derived Demographic, Sexual Behavioral, and HIV Intervention Use Measures

Using RCCS survey data, we derived demographic characteristics, sexual behavior variables, and HIV intervention use measures for index cases and controls, their partners, and female sex workers (FSWs). Demographic variables included age (grouped in 10-year categories), religion (Christian vs. non-Christian/other), community type (inland semi-urban and agrarian vs. Lake Victoria fishing), marital status (never married, currently married, previously married [separated/divorced/widowed]), educational attainment (no formal education, primary, secondary, technical/university, other), and primary occupation. Mobility indicators included whether participants had migrated into the study community within the last 18 months (in-migrant vs. long-term resident) and whether they reported spending at least one night away from home in the past 12 months.

Sexual behavior variables were derived from two complementary data sources: the RCCS survey, which includes an egocentric partnership module capturing detailed information on up to four recent partners, and sexual network contact tracing data, described below. Because contact tracing may be incomplete and did not collect detailed partnership-level behavioral data (so as to avoid inadvertent disclosure of reported partners) partnership characteristics were derived from the egocentric module. From this module, we constructed dichotomous indicators (yes/no) for the following: ≥1 age-disparate partnership (male partner ≥5 years older than female), ≥1 intergenerational partnership (male partner ≥10 years older), self-reported alcohol use before sex with ≥1 partner, and ≥1 partner who consumed alcohol before sex.

HIV intervention use variables included consistent condom use with casual partners, also derived from the egocentric partnership module. Condom use was summarized as a dichotomous measure of consistent use, defined as always using condoms with casual partners or reporting no casual partners. Additional HIV prevention variables, also coded as dichotomous measures, included self-reported male circumcision status, ever and current use of PrEP, and self-report of HIV testing through assisted partner notification programs.

### Contact Tracing Derived Sexual Network Measures

In contrast, contact tracing data were used to characterize the broader structure and composition of sexual networks. As part of contact tracing, all sexual partners in the previous 24 months were reported by index cases and controls. These data were used to create the following variables: number of reported partners, number of enrolled partners, number of reported FSW partners, number of reported female non-sex-worker partners, a dichotomous variable indicating whether the index had an enrolled partner with HIV, and a dichotomous variable indicating whether the index had an enrolled partner with HIV who was viremic (≥200 copies/ml).

### HIV Deep Sequencing and Genetic Linkage Analysis

HIV deep sequencing was done on two groups: 1) participants viremic at enrollment; and 2) participants who were suppressed at enrollment but had a blood sample with detectable virus collected during a previous RCCS visit. Deep sequencing was done on the Illumina platform using the veSEQ-HIV protocol (23). Tamura-Nei (TN93) distances were calculated for the whole genome, and separately for *gag*, *pol*, and *env*, with linkage defined at region-specific thresholds (*pol* < 2%, *gag* < 3%, or *env* < 5%). More detail is available in the Supplemental Methods.

### Statistical Analysis

We compared socio-demographic, mobility, and sexual behavioral characteristics of cases and controls and their enrolled sexual partners, stratified by sex. We assessed the characteristics of contact-tracing partner networks and estimated the prevalence of HIV, HIV viremia, and the reach of prevention interventions, overall and stratified by age. We fit conditional logistic regressions for all participants and stratified by sex. To address potential confounding, we adjusted for marital status and education of the index in sensitivity analyses.

### Transmission Modeling

Because the source of infection cannot be directly observed and many partners were untraceable, we used transmission modeling to estimate the proportion of incident cases attributable to FSW partnerships. We developed two transmission models: simple and network-based. The network-based model was a Bayesian additive cumulative hazard acquisition model with dyad-specific transmission weights derived from partner covariates, whereas the simple model assigned a random partner with HIV to be the source of infection. Together with four different imputation strategies for untraceable partners, these approaches yielded eight estimates for the proportion of transmissions originating from FSWs. A complete explanation of imputation and modeling methods is available in the Supplemental Methods.

### Ethics

This study was approved by the Uganda Virus Research Institute Research and Ethics Committee, the Uganda National Council for Science and Technology, and the Johns Hopkins School of Medicine. All Rakai Community Cohort Study (RCCS) and case-control participants provided written informed consent.

## RESULTS

### HIV Testing

A total of 34,033 censused individuals were eligible for the RCCS during the study period (Supplemental Figure 1) and 22,415 (65.9%) agreed to participate. We administered a total of 38,899 HIV tests to 22,255 of these participants over the study period and identified 3,926 (17.6%) people living with HIV.

We identified 187 (98 female, 89 male) incident HIV cases, of which 164 (85 female, 79 male) were enrolled in this nested study (Figure 1). Incident cases were predominantly identified through longitudinal HIV testing (n=136), followed by viral load testing and a self-reported previous negative serologic test (n=42), and recency and viral load testing (n=9). Unenrolled cases (n=23) were either untraceable (n=5), refused enrollment (n=10), or were only identified as eligible retrospectively after study completion (n=8).

**Figure 1:**
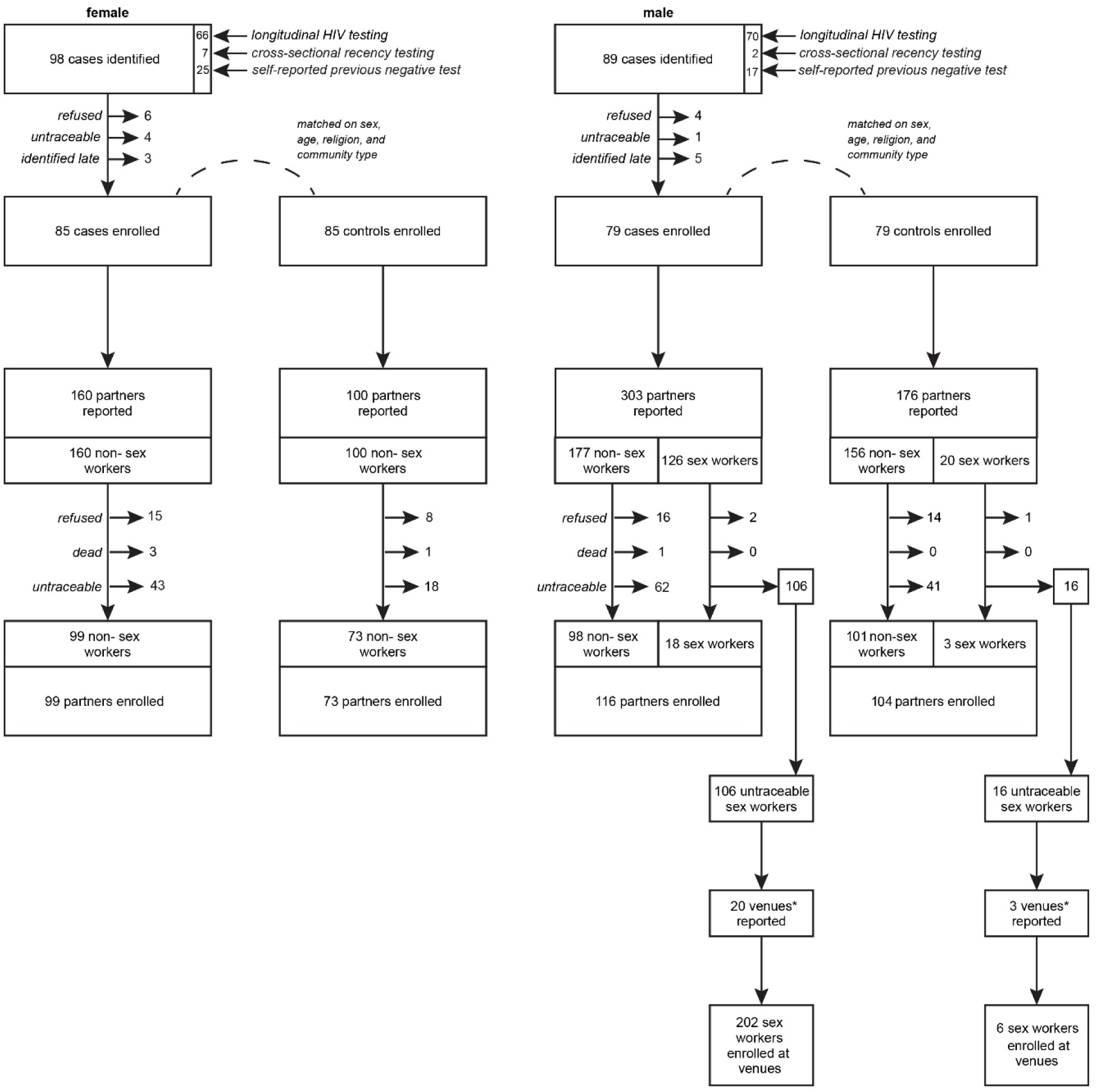
Enrollment Flow Chart. Index cases were matched to HIV-negative controls through sex, age (+/− 2 years), religion (Christian or not Christian), and community type (fishing or inland). *Counting only actively operating venues

### Index Characteristics

Case and control index participants were successfully matched across sex, age, religion, and community type. Both male (31.6%) and female (32.9%) cases were more likely to be previously, but no longer, married than male (13.9%) and female (15.3%) controls. Male cases were less likely to have only stable partners or consistently use condoms with casual partners compared to male controls (32.9% vs 51.9%). Female cases were more likely to consume alcohol before sex (39.1% vs 18.8%) or have a partner who did so (60.9% vs 39.1%) compared to female controls (Supplemental Table 1).

### Sexual Network Size and Partner Enrollment

Overall, 739 total partners were reported through contact-tracing activities: 303 (41.0%) from male cases, 176 (23.8%) from male controls, 160 (21.7%) from female cases, and 100 (13.5%) from female controls (Figure 1). A median of 3 (IQR:2-5) partners were reported from male cases, compared to 2 (1–3) for male controls, 2 (1–2) for female cases, and 1 (1–1) for female controls (Figure 2a). The difference in partners reported between male cases and controls was largely (106/127,83.5%) explained by FSW partners. Among male incident cases, those with FSW partners had substantially larger sexual networks (median 6 partners; IQR 3-8 vs 2;2-3), reflecting multiple FSW partnerships (median 3;IQR 1-5) combined with similar numbers of non-FSW partners (median 2 in cases and controls).

**Figure 2:**
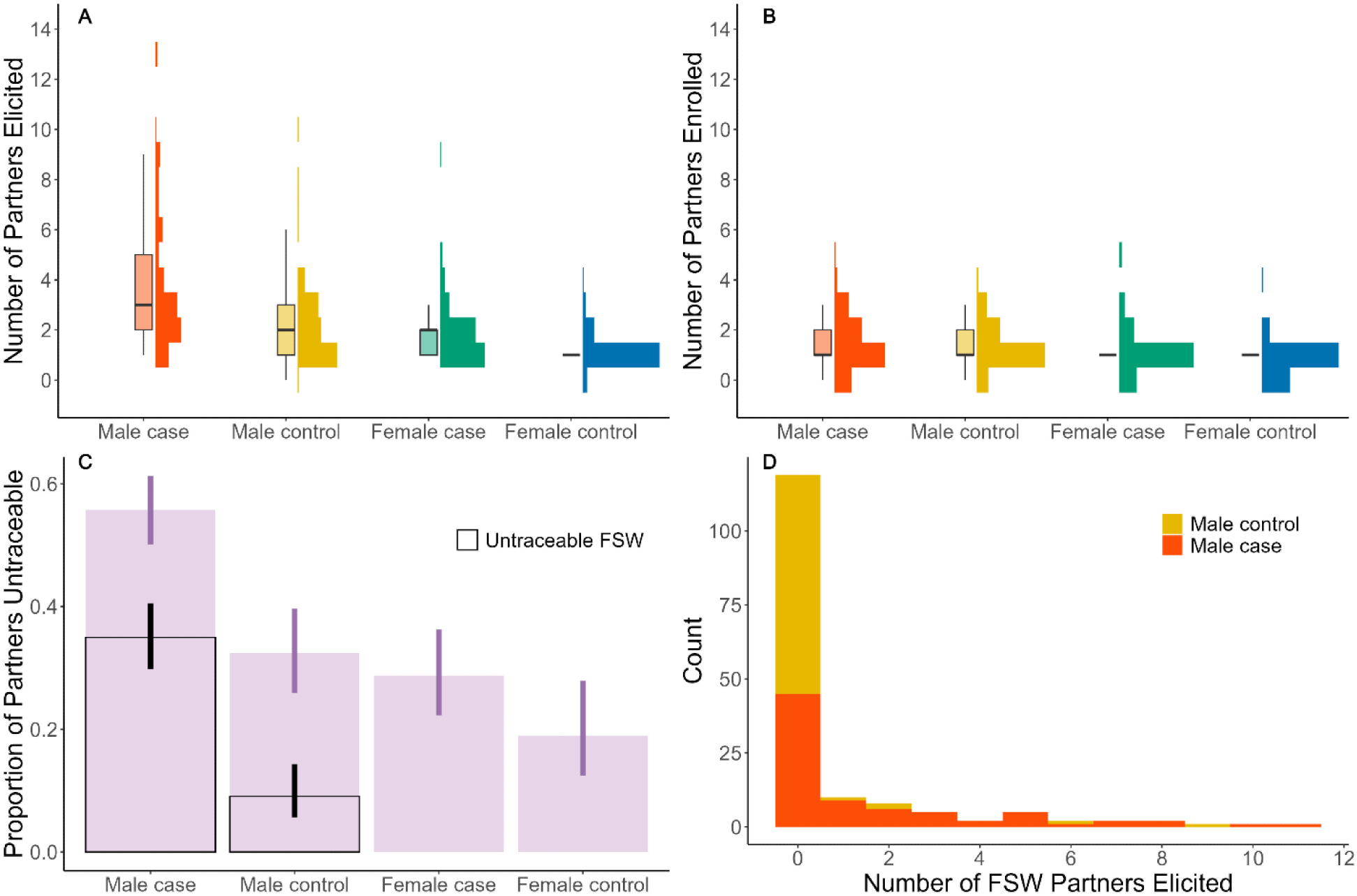
Partner Reporting and Enrollment. Shown are the distributions of the number of reported partners per-index by sex and arm (Panel A), number of enrolled partners per-index (Panel B), proportion of untraceable partners (Panel C), and number of reported FSW partners per index (Panel D). In panels A and B, each sex and arm strata includes a histogram displaying the frequency of raw counts and a box plot. In panel C, the proportion of untraceable partners is shown in purple, with an associated 95% confidence interval for the proportion in a darker purple. Similarly, the overall proportion of partners that are untraceable FSW is shown in black, with an associated 95% confidence interval for the proportion. In panel D, the number of FSW partners reported are shown in a stacked bar chart, where each bar is stratified by arm.

Of the 739 partners reported, 392 (53.0%) were enrolled (Figure 1). Enrollment was especially challenging for FSW partners (21/146,14.4%) because identifiable information outside of the venue was rarely provided; consequently, the distribution of enrolled partners was more similar between male cases and controls than the distribution of total partners (Figure 2b-d). Among partners with sufficient information for tracing, enrollment was high across groups: female cases: 99/114 (86.8%), female controls: 73/81 (90.1%), male cases: 116/134 (86.6%), male controls: 104/119 (87.4%). Overlap between partners of indexes was minimal: two female controls, and one male case and one male control, each shared an enrolled partner (Figure 3).

**Figure 3:**
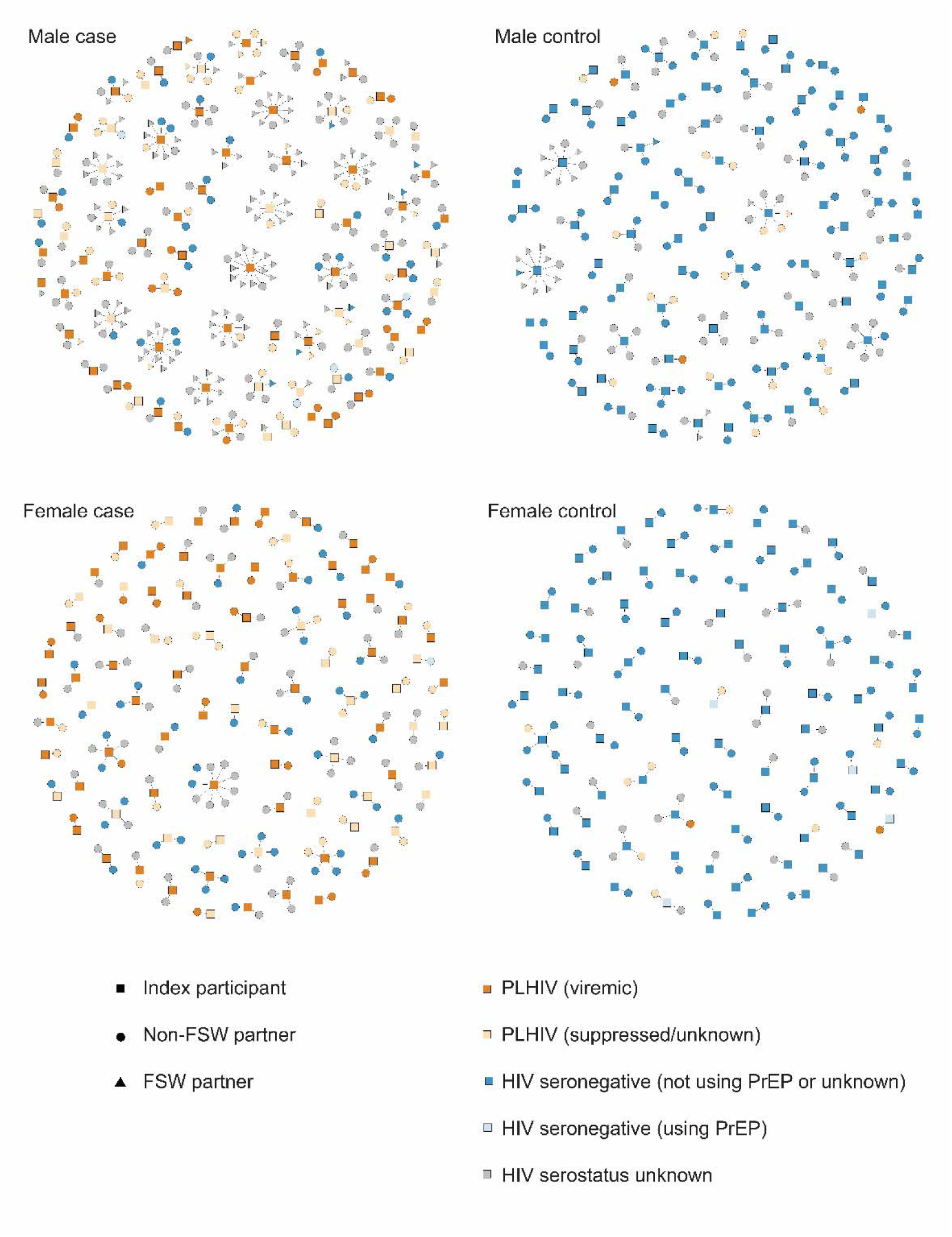
Sexual Partner Networks of Matched Cases and Controls Stratified by Index Sex. Index participants (squares) include population-representative incident HIV cases and matched controls from Southern Uganda from 2021-2024. Contact tracing linked non-sex-workers (circles) and FSW (triangles) partners to index participants. Orange shapes indicate people with HIV, with dark orange indicating HIV viremia (≥200 copies/ml) and light orange indicating viral suppression or unknown suppression status. Blue shapes indicate people without HIV, with light blue indicating current PrEP use and dark blue indicating no current PrEP use or unknown PrEP status. Gray shapes indicate unknown HIV serostatus, usually due to the partner being untraceable.

### FSWs and venues

After the protocol was modified to include venue sampling, 28 venues were reported (Supplemental Table 2). All 22 active venues were bars, guesthouses/lodges, or both, and had estimated FSW populations between 3-200. From these venues, 208 FSWs were enrolled (Supplemental Figure 2a). Venues reported from male indexes were usually <5 kilometers (50.0%) or >30 kilometers (42.3%) from their residence (Supplemental Figure 2b). Only 11.8% (n=2/17) of reported, still actively operating, venues within the regional HIV control program catchment had been previously mapped through routine key population program activities.

### Partner and FSW Characteristics

Enrolled female partners of male cases, including 18 FSWs, were more likely to be previously married (41.2% vs 27.2%) and consume alcohol before sex (43.5% vs 24.3%) than partners of male controls (including 3 FSWs). Similarly, the majority of venue-enrolled FSWs were previously married (76.4%) and reported having a partner consume alcohol before sex (88.4%). Enrolled male partners of female cases were more likely to be employed in a fishing-related occupation (33.3% vs 22.2%) and to be older than 40 (34.3% vs 26.0%) than partners of female controls (Supplemental Table 3).

### HIV Prevalence and Viremia Within Networks

HIV prevalence was higher among predominantly non-sex-worker (non-SW) enrolled female partners of male cases (59.5%) than female partners of male controls (22.8%) and venue-enrolled FSWs (33.7%). Male partners of female cases (47.4%) also had elevated HIV prevalence compared to male partners of female controls (15.5%) (Table 1). These differences were accompanied by markedly higher levels of unsuppressed viremia within case-associated networks: 15.0% and 17.9% of female and male enrolled partners of cases were viremic compared to only 3.0% and 2.8% of female and male partners of controls and 3.4% of venue-enrolled FSWs (Table 1).

**Table 1:** HIV and Prevention Interventions Across Sexual Networks.

|  | Female Partners<br>of Male Cases | Female Partners<br>of Male Controls | Venue-Enrolled<br>FSW | Male Partners of<br>Female Cases | Male Partners of<br>Female Controls |
| --- | --- | --- | --- | --- | --- |
| HIV Prevalence<br>(95% CI) | 59.5% (49.7, 68.7) | 22.8% (15.0, 32.2) | 33.7% (27.2, 40.6) | 47.4% (37.0, 57.9) | 15.5% (8.0, 26.0) |
| Viremia<br>Prevalence<br>(95% CI) | 15.0% (8.8, 23.1) | 3.0% (0.6, 8.5) | 3.4% (1.4, 7.0) | 17.9% (10.8, 27.1) | 2.8% (0.3, 9.8) |
| Viral Suppression^ | 46/62 (74.2%) | 19/22 (86.4%) | 60/67 (89.6%) | 28/45 (62.2%) | 9/11 (81.8%) |
| Consistent<br>Condom Use# | 53/113 (46.9%) | 65/97 (67.0%) | 125/206 (60.7%) | 49/93 (52.7%) | 30/68 (44.1%) |
| Circumcision | - | - | - | 66/94 (70.2%) | 49/68 (72.1%) |
| Ever PrEP Use* | 7/45 (15.6%) | 3/75 (4.0%) | 44/136 (32.4%) | 3/48 (6.3%) | 3/58 (5.2%) |
| Current PrEP Use* | 4/45 (8.9%) | 0/75 (0.0%) | 10/136 (7.4%) | 1/48 (2.1%) | 0/58 (0.0%) |
| Testing Through<br>APN | 16/112 (14.3%) | 8/99 (8.1%) | 29/206 (14.1%) | 2/93 (2.2%) | 0/68 (0.0%) |

^Among people with HIV
\*Among people without HIV unless index cases (incident infection)

### Genomic Linkage

Genomic linkage analyses demonstrated that when viremic partners were identified, they typically formed a transmission pair with the index. Among sequenced viremic index-partner pairs, genetic linkage was observed in 17/20 partnerships (85.0%), whereas linkage was rare when partners were virally suppressed (1/7;14.3%) (Supplemental Figure 3). Genome-wide analyses of 11,696 putative pairs identified very few additional plausible transmission links beyond those identified through reported partnerships (Supplemental Figure 4).

### Intervention Coverage in Case Networks

Enrolled partners of cases had the lowest HIV suppression levels of all groups (74.2% in female partners, 62.2% in male partners), consistent condom usage proportions below 50%, and current PrEP use levels below 10% (Table 1). Elevated HIV and viremia prevalences and low uptake of prevention interventions in case networks were consistent across age groups (Supplemental Figures 5a-d, 6a-f).

### Predictors of Incident HIV Infection

Reporting of at least one FSW partner was common among male cases (43.0%) compared to controls (6.3%) and was the strongest overall predictor of incident HIV (matched odds ratio: 15.5; 95% CI: 3.7,64.8) (Figure 4). Total number of reported partners was also a strong predictor of incident HIV. However, total number of female non-SW partners was a weak predictor in men, suggesting risk was largely driven by FSW partners (Supplemental Table 4, Supplemental Figure 7). Having at least one enrolled partner with HIV (11.8;95% CI:5.1,27.2), and at least one enrolled partner with HIV viremia (7.8; 2.7,22.0) were also strongly associated with incident HIV infection, with HIV infection being especially predictive in men and viremia being especially predictive in women (Supplemental Figure 7). The most important non-network predictor was being previously married (3.1;1.7,5.6), overall and for both sexes (Supplemental Table 4, Supplemental Figure 7). Other statistically significant predictors included education, circumcision, condom use, and alcohol use before sex. Adjustment for potential confounders had a minimal impact (Supplemental Table 4).

**Figure 4:**
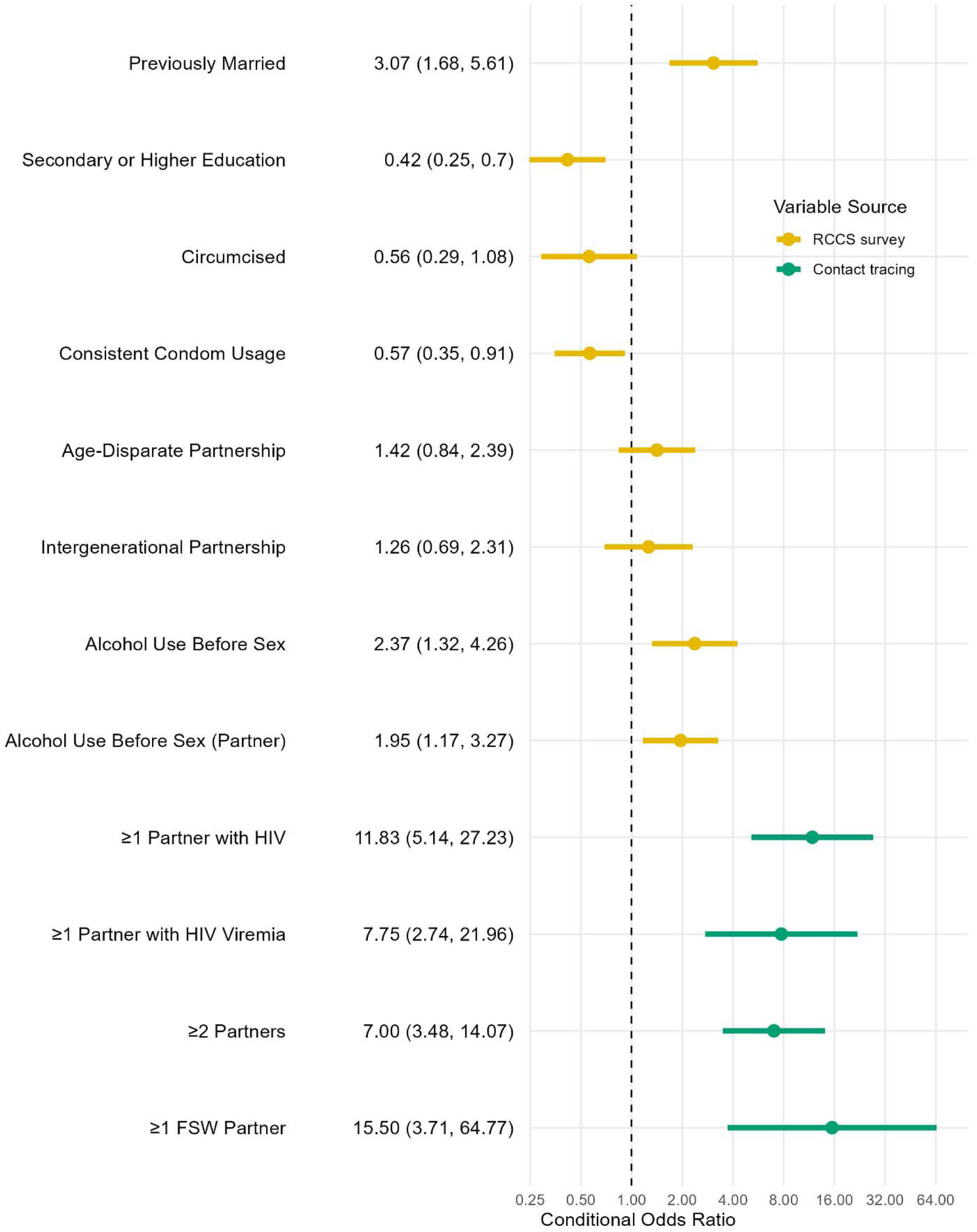
Demographic, Behavioral, and Network Predictors of Incident HIV. Points represent crude conditional odds ratio estimates, while lines represent the associated 95% confidence interval. Yellow points and intervals indicate variables that were collected during the Rakai Community Cohort Study (RCCS) survey or were constructed using those variables, while green points and intervals represent variables collected during contact tracing after enrollment in this case-control study. All variables were dichotomous, with the comparison group being the inverse of the exposed group. Previously married includes those widowed and divorced, and the comparison group includes people who are married and never married. Consistent condom usage includes those who always use condoms with casual partners or who have no casual partners. An age disparate partnership is defined as a partnership where the man was 5 or more years older than the woman and an intergenerational partnership was one where the man was 10 or more years older. Only enrolled partners could be tested for HIV and HIV viremia; however, total number of partners includes untraceable reported partners. Condom use and partner ages were obtained from partner block data from the Rakai Community Cohort Study and collapsed into dichotomous variables as described in the Supplemental Methods.

### Contribution of Sex-Work-Associated Partnerships

Men with FSW partners accounted for 43.0% of male incident infections, and 91.2% of these men also had female non-SW partners. Our mathematical model estimated that transmissions from FSW partners accounted for 16.6% (95% CI:15.2%,17.7%) of all incident infections, 34.4% (31.5%,36.8%) of male incident infections, and 80.0% (73.2%,85.4%) of incident infections in men with FSW partners. Sensitivity analyses with varying imputation and transmission assumptions estimated that between 12.9% and 18.0% of all infections were attributed to partnership with a FSW, corresponding to 26.8%-37.4% among all men, and 62.2%-86.9% among male clients (Supplemental Table 5, Supplemental Figure 8).

## DISCUSSION

In this population-representative study of incident HIV in a high-prevalence, low-incidence African epidemic (2,11,15,19), male incident cases had substantially larger sexual networks than female cases or controls of either sex. Compared to male controls, expanded male case networks were driven primarily by partnerships with FSWs, and having a FSW partner was the strongest overall predictor of incident infection, with more than four in ten male cases linked to FSWs. Most FSW partners were untraceable, as men were often only able to identify FSW venues. HIV-negative partners of cases, regardless of sex, were rarely using PrEP, indicating that individuals at high risk remain insufficiently reached by prevention services. Together, these findings suggest that ongoing transmission is disproportionately concentrated within large male client networks characterized by low uptake of prevention interventions.

Notably, this concentration of cases in FSW networks did not manifest as tightly clustered local outbreaks. Cases rarely shared reported partners or FSW venues, and genetic analyses identified few additional linkages, consistent with a locally fragmented transmission structure. A similar lack of clustering has been noted in phylogenetic studies throughout ESA (9,24–28). This consistent finding may reflect the dynamic and episodic nature of sex work, as well as geographic mobility that diffuses transmission across broader networks (29). Consistent with this fragmented structure, reported local venues were typically small and not included on existing key population program venue lists, suggesting gaps in routine outreach. Incorporating systematic venue reporting into assisted partner notification programs for newly diagnosed men, which is not currently standard of care, could support targeted prevention efforts in areas of active transmission.

Prior studies support a prominent role for male clients of FSWs and men more generally in population-level HIV transmission in African settings. For example, the Key-Pop model (30) estimated that eliminating transmissions during commercial and non-commercial sexual partnerships involving male clients would have reduced overall HIV infections by 6.9% and 41.9%, respectively, from 2010-2019. Furthermore, phylogenetic studies show that men are disproportionately and increasingly driving new HIV infections as overall HIV incidence declines, contributing to a widening female to male HIV incidence ratio across Africa (10,11). These prior findings are aligned with our results showing an outsized role of key populations and their bridging networks.

Despite their central role in epidemic control, key populations have experienced substantial and disproportionate reductions in HIV prevention funding since 2025, in the context of wider global health financing cuts (31,32). Notably, male clients of FSWs are not recognized as a formal key population in UNAIDS or PEPFAR guidance (33,34) and remain understudied as candidates for long-acting injectable PrEP and other targeted prevention strategies (35). This is particularly relevant given that African men have lower HIV diagnosis and treatment coverage levels (1,36) and are more likely to remain persistently viremic than women (11,37,38). Overall, our findings suggest continued investment in prevention programs for FSWs, alongside targeted engagement of male clients, will be critical to maintain epidemic control.

Marital dissolution also emerged as an important correlate of incident infection. Being previously, but no longer, married was strongly associated with incident HIV, and three-quarters of venue-enrolled FSWs were previously married, suggesting that marital dissolution may serve as a pathway towards HIV acquisition and entry into sex work. A recent study in Zambia found that being recently divorced or widowed was predictive of both new HIV risk behaviors and HIV acquisition (39). Other studies have found that divorced and widowed women represent large proportions of female sex workers in ESA (40), while divorced men may be more likely to engage in transactional sex (41).

This study has limitations. Our data were limited to southern Uganda, but HIV epidemiological trends in the RCCS, including declining incidence (2), fragmented transmission structures (9), and disproportionate onward male transmission (11), are consistent with many other late-stage African epidemics (2,10). We were also unable to locate a proportion of reported partners, particularly FSW partners. However, enrollment exceeded 85% across traceable partners regardless of sex and study arm, comparing favorably with APN programs in the region (42–47). Viral suppression was common at the time of sampling, and thus genetic linkages between partners could often not be assessed. However, when genetic linkages could be evaluated, viremic partners were almost always linked to the index. Our transmission models used imputation to account for untraceable partners and allowed virally suppressed partners to be considered as potential sources of infection, recognizing that transmission could have occurred before viral suppression. However, inferred transmission pathways involving untraceable and suppressed individuals should be interpreted with caution. We cannot rule out the possibility that recall bias contributed to the observed association between self-reported FSW partners and HIV incidence. However, if recall bias were the primary driver, one would expect similarly strong associations for other self-reported sexual risk behaviors, including the number of non-sex-worker partners, but these variables displayed low to modest associations with HIV incidence. Finally, sex work client status was not collected for male partners of female index cases, precluding identification of male client partnerships as a correlate of female incident infection.

In summary, our results from this novel case-control, contact-tracing study in a declining African HIV epidemic suggest diffuse transmission networks, with male clients of FSWs disproportionately represented among incident infections. These men had large sexual networks with very low PrEP coverage, suggesting high risk of onward transmission. Increased prioritization of male clients of FSWs, including for long-acting injectable PrEP, could be a highly efficient strategy for HIV prevention and epidemic control in Africa.

## Supporting information

Supplementary Methodology

## Data Availability

All deidentified data produced in the present study are available upon reasonable request to the authors.

## ACKNOWLEDGEMENT

MKG and LWC were supported by NIH grant R01AI143333. GJB was supported by NIH grant T32AI102623. TG is supported by an Investigator Grant (GNT2025445) from the National Health and Medical Research Council, Australia (NHMRC). JL was supported by the University of North Carolina at Chapel Hill Center for AIDS Research (NIH P30AI050410). AK was supported by NIH grant F31MH134699. This research was supported [in part] by the Intramural Research Program of the National Institutes of Health (NIH). The contributions of the NIH author(s) are considered Works of the United States Government. The findings and conclusions presented in this paper are those of the author(s) and do not necessarily reflect the views of the NIH or the U.S. Department of Health and Human Services. Whole-genome sequences were generated by PANGEA-HIV consortium, through funding by the Bill and Melinda Gates Foundation (OPP1084362, INV-007573, INV-060259 and INV-075093).

## Notes

### Competing Interest Statement

The authors have declared no competing interest.

