## Supplementary Appendix for "HIV Transmission in a Declining African Epidemic"

**Table of Contents**

Supplemental Figure 1: Identification of Incident HIV Infections Among Rakai Community Cohort Study Participants	2

Supplemental Table 1: Characteristics of Index Participants, Stratified by Study Group and Sex	3

Supplemental Table 2: Characteristics of Reported Venues	5

Supplemental Figure 2: Enrollment from Female Sex Worker Venues, and Distance Between Index Participant Residential Locations and Linked Venues	6

Supplemental Table 3: Characteristics of Enrolled Partners and Sex Workers, Stratified by Study Group and Sex	7

Supplemental Figure 3: Distribution of Genetic Distances Among HIV Sequence Pairs	9

Supplemental Figure 4: HIV Transmission Networks Integrating Contact Tracing and Genetic Data	10

Supplemental Figure 5: HIV Prevalence and Viremia Among Sexual Partners of Incident HIV Cases, Matched Controls, and Female Sex Workers	11

Supplemental Figure 6: Distribution of HIV Viral Suppression, PrEP Use, and Assisted Partner Notification Testing Among Sexual Partners of Incident HIV Cases and Controls	12

Supplemental Table 4: Crude and Adjusted Matched Case-Control Odds Ratios from Conditional Logistic Regression, Overall and Stratified by Sex	14

Supplemental Figure 7: Demographic, Behavioral, and Network Predictors of Incident HIV, Stratified by Sex	16

Supplemental Table 5: Transmission Model Main and Sensitivity Analysis Results	17

Supplemental Figure 8: Transmission Model Main and Sensitivity Analysis Posterior Distributions	18

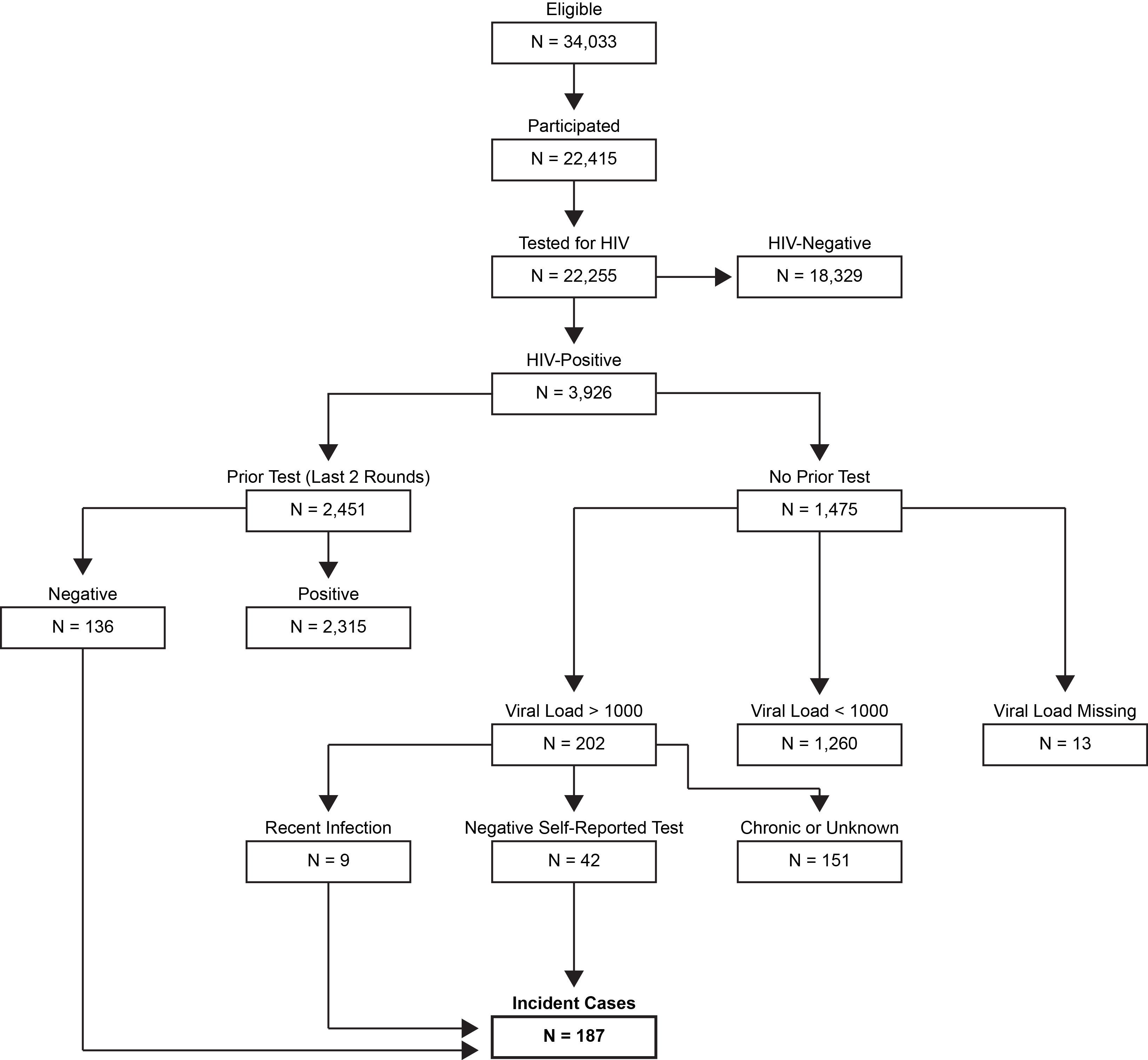

### Supplemental Figure 1: Identification of Incident HIV Infections Among Rakai Community Cohort Study Participants

*Flow diagram illustrating the selection of incident HIV cases from participants in the Rakai Community Cohort Study (RCCS). Among 34,033 eligible individuals, 22,415 participated in the RCCS, of whom 22,255 underwent HIV testing. Participants who tested HIV-seronegative (N = 18,329) were excluded from further classification. Among those testing HIV-seropositive (N = 3,926), prior HIV testing history and viral load measurements were used to classify infection status. Individuals with a documented negative HIV test within the two preceding survey rounds who newly tested positive were classified as incident infections. Among individuals without a prior documented test, viral load and self-reported testing history were used to distinguish recent infections from chronic or previously diagnosed infections. Participants with a viral load >1000 copies/mL and a negative self-reported prior test were classified as incident infections. The final analytic sample included 187 incident HIV cases.*

### Supplemental Table 1: Characteristics of Index Participants, Stratified by Study Group and Sex

|  | **Male Cases  N = 79** | **Male Controls N = 79** | **Female Cases N = 85** | **Female Controls N = 85** |
| --- | --- | --- | --- | --- |
| *Demographics* |  |  |  |  |
| **Age (years)** |  |  |  |  |
| Median (IQR) | 31 (25.5, 37) | 31 (25, 37) | 31 (25, 38) | 31 (25, 38) |
| 18-29 | 36 (45.6%) | 35 (43.0%) | 35 (41.2%) | 37 (43.5%) |
| 30-39 | 29 (35.4%) | 31 (39.2%) | 33 (38.8%) | 32 (37.6%) |
| 40-49 | 12 (15.2%) | 11 (13.9%) | 14 (16.5%) | 13 (15.3%) |
| 50-59 | 3 (3.8%) | 3 (3.8%) | 3 (3.5%) | 3 (3.5%) |
| **Religion** |  |  |  |  |
| Christian | 69 (87.2%) | 69 (87.2%) | 72 (84.7%) | 72 (84.7%) |
| Other or none | 10 (12.8%) | 10 (12.8%) | 13 (15.3%) | 13 (15.3%) |
| **Community type** |  |  |  |  |
| Inland | 63 (79.7%) | 63 (79.7%) | 67 (78.8%) | 67 (78.8%) |
| Fishing | 16 (20.3%) | 16 (20.3%) | 18 (21.2%) | 18 (21.2%) |
| **Marital status** |  |  |  |  |
| Currently married | 45 (57.0%) | 56 (70.9%) | 52 (61.2%) | 67 (78.8%) |
| Previously married | 25 (31.6%) | 11 (13.9%) | 28 (32.9%) | 13 (15.3%) |
| Never married | 9 (11.4%) | 12 (15.2%) | 5 (5.9%) | 5 (5.9%) |
| **Educational attainment** |  |  |  |  |
| No formal education | 3 (3.8%) | 2 (2.5%) | 8 (9.4%) | 4 (4.7%) |
| Primary | 54 (68.4%) | 45 (57.0%) | 53 (62.4%) | 39 (45.9%) |
| Secondary | 18 (22.8%) | 17 (21.5%) | 15 (17.6%) | 22 (25.9%) |
| Technical/University | 4 (5.1%) | 15 (19.0%) | 9 (10.6%) | 20 (23.5%) |
| **Primary occupation** |  |  |  |  |
| Agriculture or Housework | 18 (22.8%) | 19 (24.1%) | 44 (51.8%) | 42 (49.4%) |
| Trading or Shopkeeping | 19 (24.1%) | 12 (15.2%) | 17 (20.0%) | 24 (28.2%) |
| Bar or restaurant work | 0 (0.0%) | 0 (0.0%) | 12 (14.1%) | 10 (11.8%) |
| Sex work | 0 (0.0%) | 0 (0.0%) | 0 (0.0%) | 0 (0.0%) |
| Fishing-related occupation | 22 (27.8%) | 26 (32.9%) | 12 (14.1%) | 9 (10.6%) |
| Other | 20 (25.3%) | 22 (27.8%) | 44 (51.8%) | 42 (49.4%) |
| *Mobility*  **Migration** |  |  |  |  |
| In-migrant | 29 (37.2%) | 29 (37.2%) | 58 (69.0%) | 44 (52.4%) |
| Long-term resident (18+ months) | 49 (62.8%) | 49 (62.8%) | 26 (31.0%) | 40 (47.6%) |
| *Missing* | 1 | 1 | 1 | 1 |
| **Slept away from home (last 12 months)** |  |  |  |  |
| Yes | 29 (37.7%) | 34 (44.2%) | 43 (52.4%) | 41 (50.0%) |
| *Missing* | 2 | 2 | 3 | 3 |
| *Sexual Behavior (RCCS^)* |  |  |  |  |
| **Number of sex partners (last 12 months)** |  |  |  |  |
| *0* | 2 (2.6%) | 2 (2.6%) | 2 (2.6%) | 2 (2.6%) |
| *1* | 23 (29.9%) | 33 (42.9%) | 23 (29.9%) | 33 (42.9%) |
| *2* | 18 (23.4%) | 20 (26.0%) | 18 (23.4%) | 20 (26.0%) |
| *3+* | 34 (44.2%) | 22 (28.6%) | 34 (44.2%) | 22 (28.6%) |
| *Missing or unknown* | 2 | 2 | 3 | 3 |
| **≥1 age-disparate sexual partnership (male ≥5 years older)** |  |  |  |  |
| Yes | 47 (62.7%) | 39 (52.7%) | 40 (54.8%) | 37 (50.0%) |
| *Missing or unknown* | 4 | 5 | 12 | 11 |
| **≥1 intergenerational sexual partnership (male ≥10 years older)** |  |  |  |  |
| Yes | 22 (29.3%) | 15 (20.5%) | 14 (19.2%) | 15 (20.5%) |
| *Missing or unknown* | 4 | 6 | 12 | 12 |
| **Alcohol consumption before sex with ≥1 partner** |  |  |  |  |
| Yes | 34 (58.6%) | 26 (44.8%) | 27 (39.1%) | 13 (18.8%) |
| *Missing* | 21 | 21 | 16 | 16 |
| **≥1 partner who consumed alcohol before sex** |  |  |  |  |
| Yes | 28 (48.3%) | 22 (37.9%) | 42 (60.9%) | 27 (39.1%) |
| *Missing* | 21 | 21 | 16 | 16 |
| *Intervention Uptake* |  |  |  |  |
| **Consistent Condom Use*** |  |  |  |  |
| *Yes* | 25 (32.9%) | 40 (51.9%) | 52 (65.8%) | 57 (73.1%) |
| *Missing* | 3 | 2 | 6 | 7 |
| **Circumcised (males only)** |  |  |  |  |
| *Yes* | 42 (54.5%) | 53 (68.8%) | - | - |
| *Missing* | 2 | 2 | - | - |
| **Ever PrEP Use** |  |  |  |  |
| *Yes* | 4 (5.1%) | 2 (2.6%) | 7 (8.4%) | 6 (7.2%) |
| *Missing* | 1 | 1 | 2 | 2 |
| **Testing Through APN** |  |  |  |  |
| *Yes* | 1 (1.3%) | 0 (0.0%) | 4 (4.8%) | 1 (1.2%) |
| *Missing* | 1 | 1 | 2 | 2 |

*Only stable partners or consistent condom use with casual partners

^ Sexual behavior variables were derived from partner blocks in the RCCS, as described in the Supplemental Methods

### Supplemental Table 2: Characteristics of Reported Venues

| **Venue Number** | **# FSW enrolled** | **Venue category** | **City (if outside study area)** | **Previously known if local and open** | **# of cases that reported the venue** | **# controls that reported the venue** | **# FSW at busy time** |
| --- | --- | --- | --- | --- | --- | --- | --- |
| 1 | 12 | Bar, guesthouse |  |  | 1 | 0 | 25 |
| 2 | 13 | Bar, guesthouse |  |  | 1 | 0 | 18 |
| 3 | 9 | Bar, guesthouse |  | Y | 1 | 0 | 14 |
| 4 | 31 | Guesthouse | Kampala | n/a | 1 | 0 | 80 |
| 5 | 2 | Bar, guesthouse |  |  | 1 | 0 | 5 |
| 7/8* | 23 | Brothel, bar, guesthouse |  | Y | 2 | 1 | 29 |
| 9 | 34 | Nightclub, bar, massage parlor, brothel, hotel | Kampala | n/a | 1 | 0 | 200 |
| 13 | 6 | Bar, guesthouse |  |  | 0 | 1 | 7 |
| 14 | 0 | Bar, guesthouse |  |  | 0 | 1 | 9 |
| 17 | 2 | Guesthouse | Mbarara | n/a | 1 | 0 | 5 |
| 18 | 4 | Nightclub, bar, brothel, guest house | Mbarara | n/a | 1 | 0 | 25 |
| 19 | 38 | Bar, guesthouse | Kampala | n/a | 1 | 0 | 90 |
| 20 | 0 | Bar, guesthouse |  |  | 1 | 0 | 4 |
| 21 | 2 | Bar |  |  | 2 | 0 | 6 |
| 22 | 0 | Guesthouse |  |  | 1 | 0 | 3 |
| 23 | 10 | Guesthouse |  |  | 1 | 0 | 20 |
| 24 | 3 | Guesthouse |  |  | 1 | 0 | 5 |
| 25 | 4 | Guesthouse |  |  | 1 | 0 | 15 |
| 26 | 9 | Nightclub, bar, brothel, guesthouse |  |  | 1 | 0 | 25 |
| 27 | 2 | Bar |  |  | 1 | 0 | 5 |
| 28 | 3 | Bar |  |  | 1 | 0 | 8 |
| 29 | 1 | Bar, guesthouse |  |  | 1 | 0 | 5 |
| c1 | 0 | Closed |  | n/a | 1 | 0 | n/a |
| c2 | 0 | Closed | Kampala | n/a | 1 | 0 | n/a |
| c3 | 0 | Closed | Kampala | n/a | 1 | 0 | n/a |
| c4 | 0 | Closed |  | n/a | 1 | 0 | n/a |
| c5 | 0 | Closed |  | n/a | 1 | 0 | n/a |
| c6 | 0 | Closed |  | n/a | 0 | 1 | n/a |

**Venues 7 and 8 are located next to each other and many FSW work both venues.*

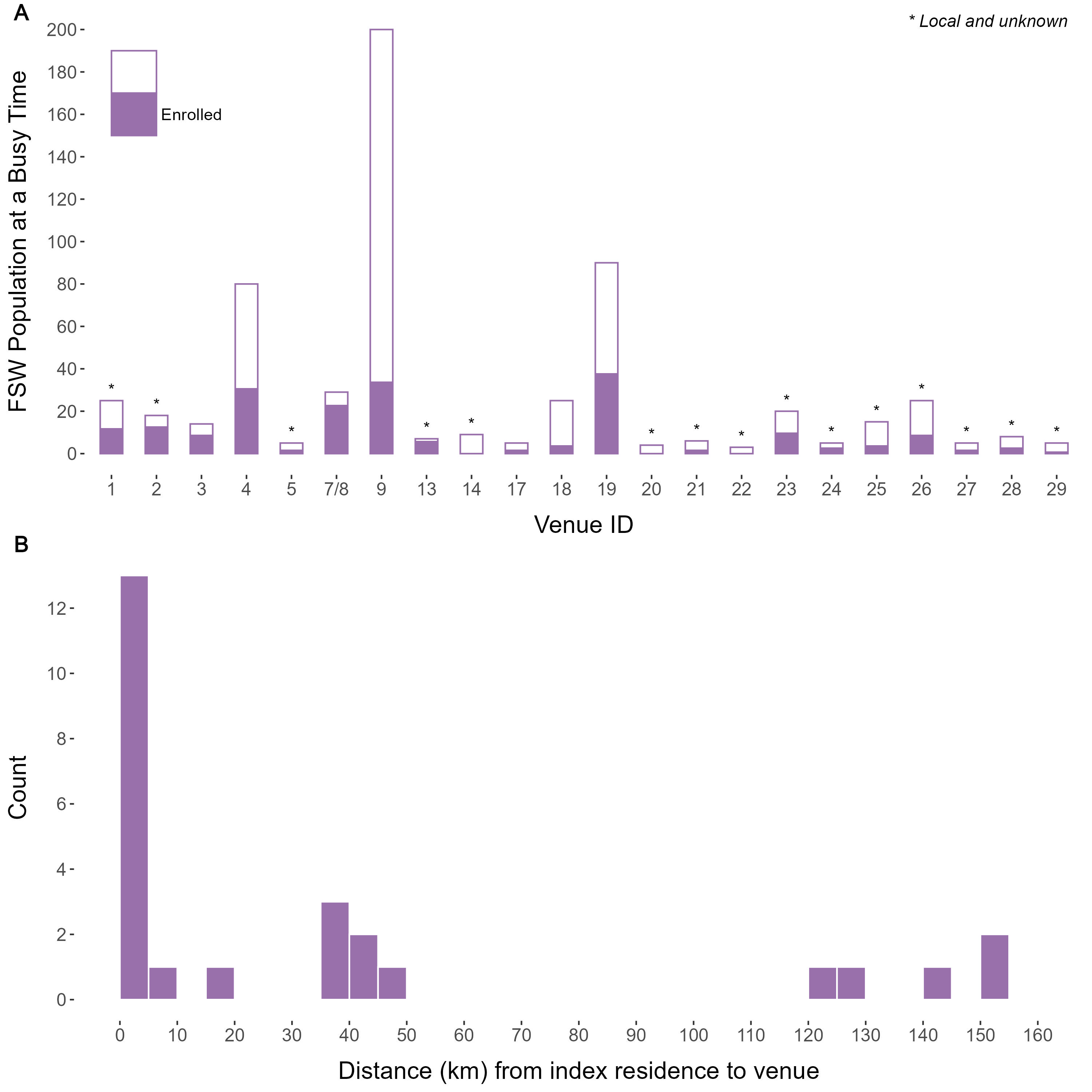

### Supplemental Figure 2: Enrollment from Female Sex Worker Venues, and Distance Between Index Participant Residential Locations and Linked Venues

*Panel (a) shows the estimated number of female sex workers (FSW) present at busy times across study venues. Busy time was defined as the period of peak activity at a venue when the largest number of FSW and clients are present, based on venue mapping and key informant reports. Empty bars indicate the estimated FSW population at each venue during busy time, and filled bars indicate the number of FSW enrolled from each venue in the study. Venues marked with an asterisk are those within the study area that were previously unknown to local HIV control programs. Panel (b) shows the distribution of distances between the residential location of index participants and the venues where linked FSW were identified.*

### Supplemental Table 3: Characteristics of Enrolled Partners and Sex Workers, Stratified by Study Group and Sex

|  | **Female Partners of Male Cases**  **N = 116** | **Female Partners of Male Controls**  **N = 104** | **Venue-Enrolled Female Sex Workers**  **N = 208** | **Male Partners of Female Cases**  **N = 99** | **Male Partners of Female Controls**  **N = 73** |
| --- | --- | --- | --- | --- | --- |
| *Demographics* |  |  |  |  |  |
| **Age (years)** |  |  |  |  |  |
| Median (IQR) | 28 (23, 34) | 28 (23, 35) | 28.5 (23, 35) | 35 (29, 42) | 35 (29, 41) |
| 18-29 | 61 (52.6%) | 56 (53.8%) | 114 (54.8%) | 27 (27.3%) | 19 (26.0%) |
| 30-39 | 38 (32.8%) | 35 (33.7%) | 75 (36.1%) | 38 (38.4%) | 35 (47.9%) |
| 40-49 | 16 (13.8%) | 10 (9.6%) | 19 (9.1%) | 29 (29.3%) | 13 (17.8%) |
| 50-59 | 1 (0.9%) | 3 (2.9%) | 0 (0.0%) | 5 (5.1%) | 6 (8.2%) |
| **Religion** |  |  |  |  |  |
| Christian | 105 (92.1%) | 81 (78.6%) | 155 (75.6%) | 77 (79.4%) | 54 (75.0%) |
| Other or none | 9 (7.9%) | 22 (21.4%) | 50 (24.4%) | 20 (20.6%) | 18 (25.0%) |
| *Missing* | 2 | 1 | 3 | 2 | 1 |
| **Community type** |  |  |  |  |  |
| Inland | 99 (85.3%) | 83 (79.8%) | 194 (93.3%) | 81 (81.8%) | 57 (78.1%) |
| Fishing | 17 (14.7%) | 21 (20.2%) | 14 (6.7%) | 18 (18.2%) | 16 (21.9%) |
| **Marital status** |  |  |  |  |  |
| Currently married | 54 (47.4%) | 67 (65.0%) | 18 (8.7%) | 73 (74.5%) | 64 (87.7%) |
| Previously married | 47 (41.2%) | 28 (27.2%) | 159 (76.4%) | 20 (20.4%) | 9 (12.3%) |
| Never married | 13 (11.4%) | 8 (7.8%) | 31 (14.9%) | 5 (5.1%) | 0 (0.0%) |
| *Missing* | 2 | 1 | 0 | 1 | 0 |
| **Educational attainment** |  |  |  |  |  |
| No formal education | 5 (4.4%) | 5 (4.9%) | 6 (2.9%) | 3 (3.0%) | 2 (2.8%) |
| Primary | 72 (63.2%) | 56 (54.4%) | 114 (55.3%) | 71 (71.7%) | 45 (62.5%) |
| Secondary | 28 (24.6%) | 23 (22.3%) | 71 (34.5%) | 17 (17.2%) | 15 (20.8%) |
| Technical/University | 9 (7.9%) | 19 (18.4%) | 15 (7.3%) | 8 (8.1%) | 10 (13.9%) |
| *Missing* | 2 | 1 | 2 | 0 | 1 |
| **Primary occupation** |  |  |  |  |  |
| Agriculture or Housework | 52 (45.6%) | 44 (42.7%) | 7 (3.4%) | 26 (26.3%) | 19 (26.4%) |
| Trading or Shopkeeping | 31 (27.2%) | 28 (27.2%) | 10 (4.9%) | 16 (16.2%) | 16 (22.2%) |
| Bar or restaurant work | 20 (17.5%) | 14 (13.6%) | 37 (18.0%) | 1 (1.0%) | 1 (1.4%) |
| Sex work | 4 (3.5%) | 1 (1.0%) | 142 (68.9%) | 0 (0.0%) | 0 (0.0%) |
| Fishing-related occupation | 0 (0.0%) | 0 (0.0%) | 0 (0.0%) | 33 (33.3%) | 16 (22.2%) |
| Other | 7 (6.1%) | 16 (15.5%) | 10 (4.9%) | 23 (23.2%) | 20 (27.8%) |
| *Missing* | 2 | 1 | 2 | 0 | 1 |
| *Mobility* |  |  |  |  |  |
| **Migration** |  |  |  |  |  |
| In-migrant | 65 (57.0%) | 57 (55.3%) | 118 (57.6%) | 36 (37.1%) | 25 (34.7%) |
| Long-term resident (18+ months) | 49 (43.0%) | 46 (44.7%) | 87 (42.4%) | 61 (62.9%) | 47 (65.3%) |
| *Missing* | 2 | 1 | 3 | 2 | 1 |
| **Slept away from home (last 12 months)** |  |  |  |  |  |
| Yes | 69 (61.1%) | 61 (61.0%) | 176 (89.6%) | 45 (47.9%) | 33 (48.5%) |
| *Missing* | 3 | 4 | 2 | 5 | 5 |
| *Sexual Behavior (RCCS^)* |  |  |  |  |  |
| **Number of sex partners (last 12 months)** |  |  |  |  |  |
| *0* | 1 (0.9%) | 3 (3.0%) | 0 (0.0%) | 2 (2.1%) | 0 (0.0%) |
| *1* | 64 (56.6%) | 69 (69.0%) | 6 (2.9%) | 29 (30.9%) | 22 (32.4%) |
| *2* | 19 (16.8%) | 21 (21.0%) | 8 (3.9%) | 39 (41.5%) | 17 (25.0%) |
| *3+* | 29 (25.7%) | 7 (7.0%) | 192 (93.2%) | 24 (25.5%) | 29 (42.6%) |
| *Missing or unknown* | 3 | 4 | 2 | 5 | 5 |
| **≥1 age-disparate sexual partnership (male ≥5 years older)** |  |  |  |  |  |
| Yes | 66 (60.6%) | 50 (53.8%) | 148 (81.8%) | 53 (58.2%) | 46 (67.6%) |
| *Missing or unknown* | 7 | 11 | 27 | 8 | 5 |
| **≥1 intergenerational sexual partnership (male ≥10 years older)** |  |  |  |  |  |
| Yes | 37 (34.3%) | 26 (28.0%) | 105 (62.1%) | 26 (28.9%) | 24 (35.8%) |
| *Missing or unknown* | 8 | 11 | 39 | 9 | 6 |
| **Alcohol consumption before sex with ≥1 partner** |  |  |  |  |  |
| Yes | 37 (43.5%) | 18 (24.3%) | 105 (60.0%) | 32 (42.1%) | 21 (37.5%) |
| *Missing* | 31 | 30 | 33 | 23 | 17 |
| **≥1 partner who consumed alcohol before sex** |  |  |  |  |  |
| Yes | 48 (56.5%) | 37 (50.0%) | 153 (88.4%) | 29 (38.2%) | 17 (30.9%) |
| *Missing* | 31 | 30 | 35 | 23 | 18 |

^ Sexual behavior variables were derived from partner blocks in the RCCS, as described in the Supplemental Methods

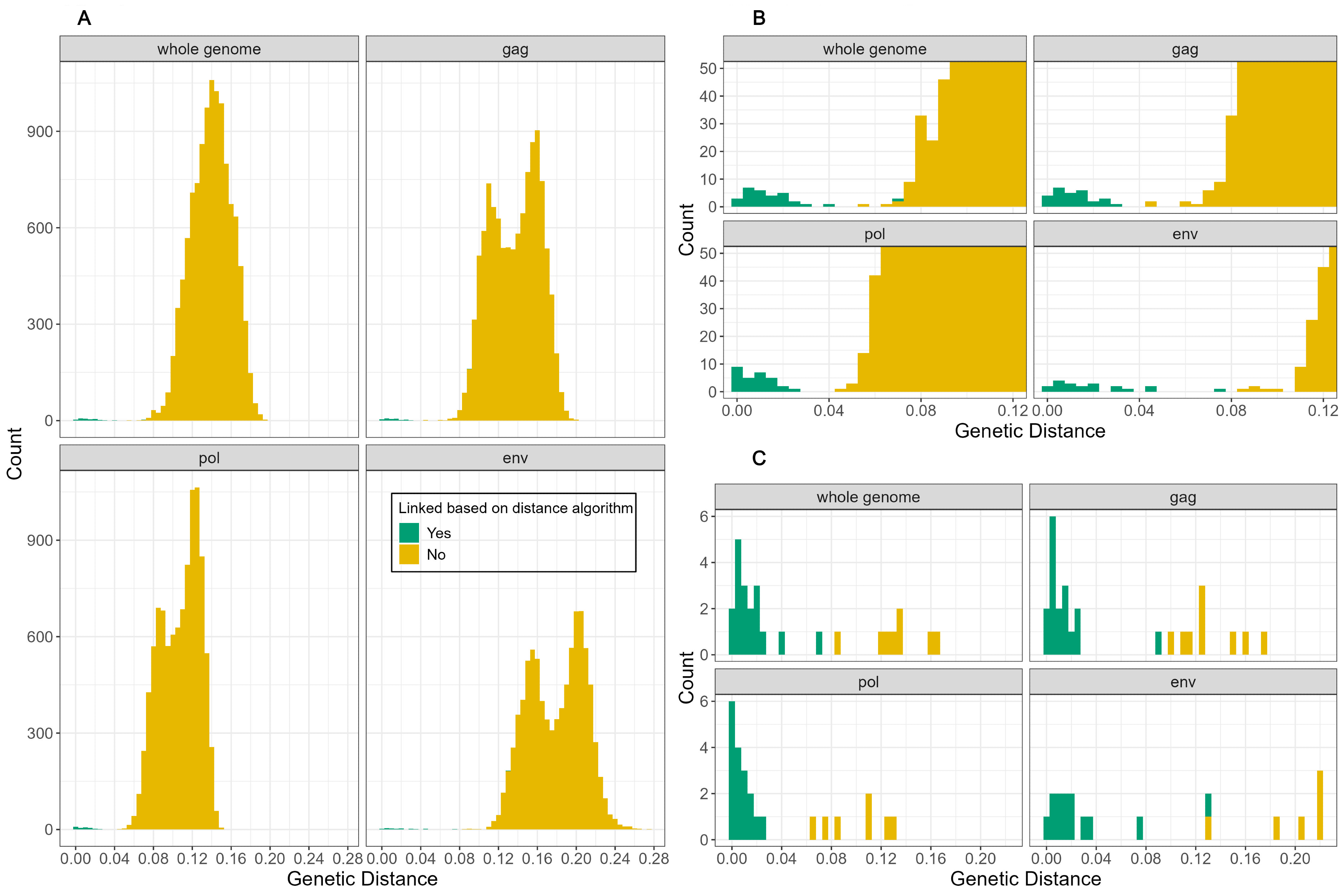

### Supplemental Figure 3: Distribution of Genetic Distances Among HIV Sequence Pairs

*Histograms showing the distribution of pairwise genetic distances across HIV sequences among participants with detectable viremia (viral load >1,000 copies/mL) or historical samples with detectable virus. Distances are shown for the whole genome and for the gag, pol, and env regions. Panel a) displays genetic distances for all possible sequence pairs in the dataset, while b) highlights the lower range of distances where putative transmission pairs are expected. Bars are colored according to whether pairs were classified as genetically linked based on the distance threshold algorithm (green) or not linked (yellow). Panel c) shows the subset of sequence pairs corresponding to partnerships identified through contact tracing.*

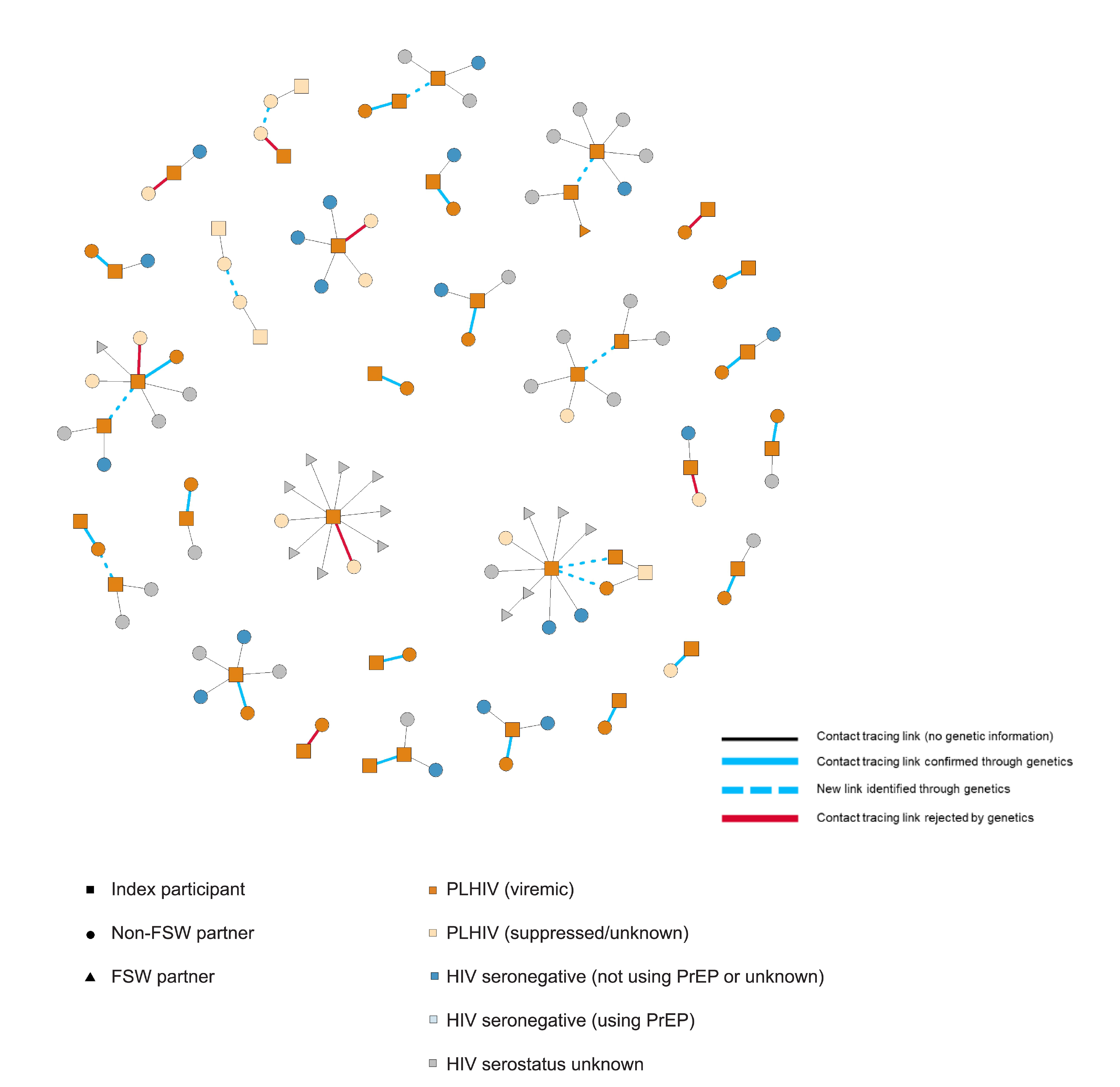

### Supplemental Figure 4: HIV Transmission Networks Integrating Contact Tracing and Genetic Data

*Network diagram illustrating sexual partnerships identified through contact tracing among participants with incident HIV infection and their partners in the Rakai Community Cohort Study. Squares represent index participants, circles represent non–female sex worker (FSW) partners, and triangles represent FSW partners. Node colors indicate HIV status and treatment status: people living with HIV (PLHIV) with viremia, PLHIV who are virally suppressed or with unknown viral load, HIV-seronegative individuals not using PrEP or with unknown PrEP use, HIV-seronegative individuals using PrEP, and individuals with unknown HIV serostatus. Solid black lines indicate contact tracing links without genetic information. Blue solid lines indicate partnerships confirmed through phylogenetic linkage. Blue dashed lines indicate additional transmission links identified through genetic data but not reported through contact tracing. Red lines indicate reported partnerships that were not identified as genetically-linked pairs.*

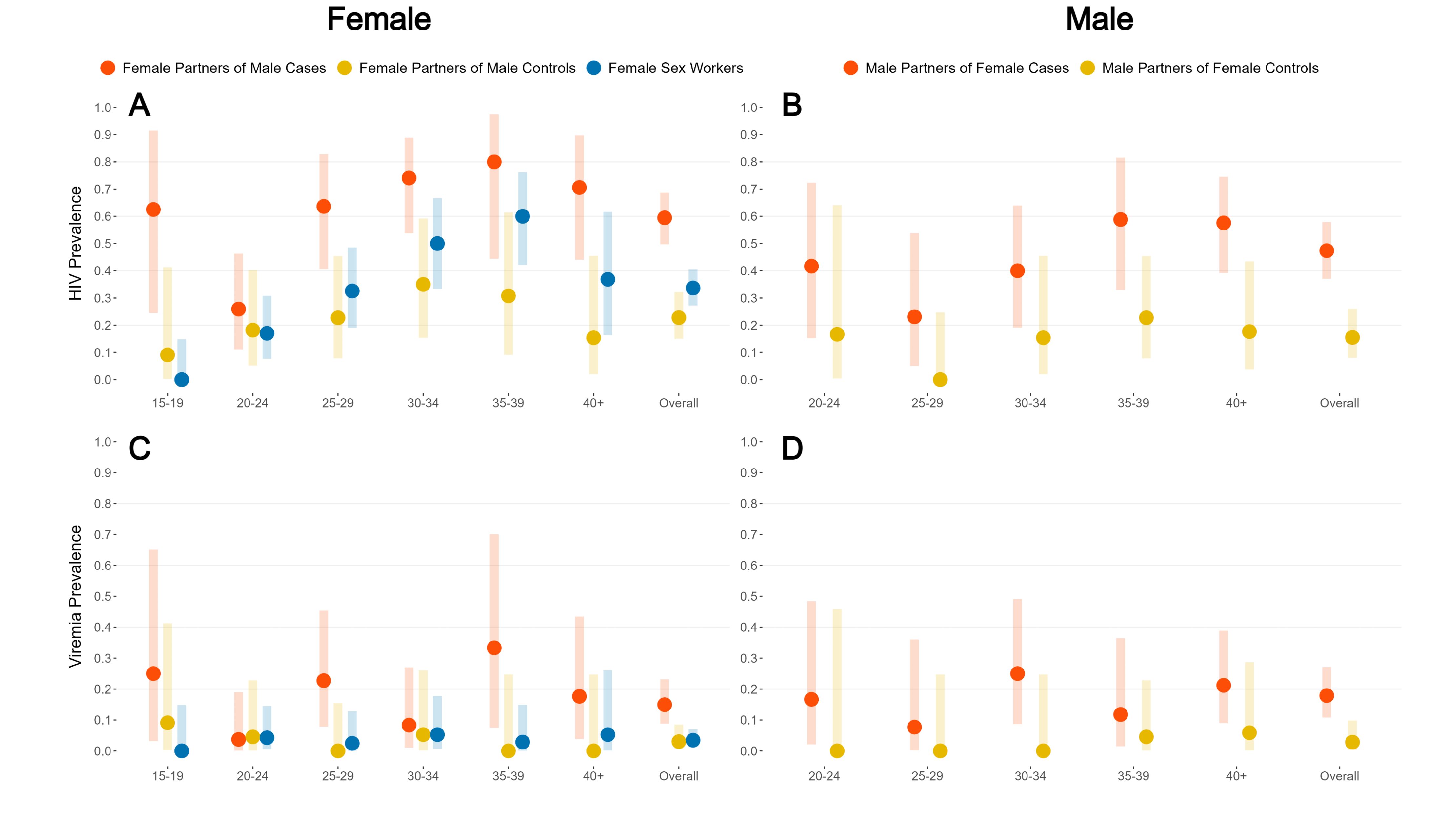

### Supplemental Figure 5: HIV Prevalence and Viremia Among Sexual Partners of Incident HIV Cases, Matched Controls, and Female Sex Workers

*Panels A and B show HIV prevalence among female partners of male indexes and FSWs (A) and male partners of female indexes (B). Panels C and D show the prevalence of viremia (viral load >1,000 copies/mL) among the same groups. Points represent prevalence estimates and bars represent 95% confidence intervals. Results are shown overall and stratified by age group.*

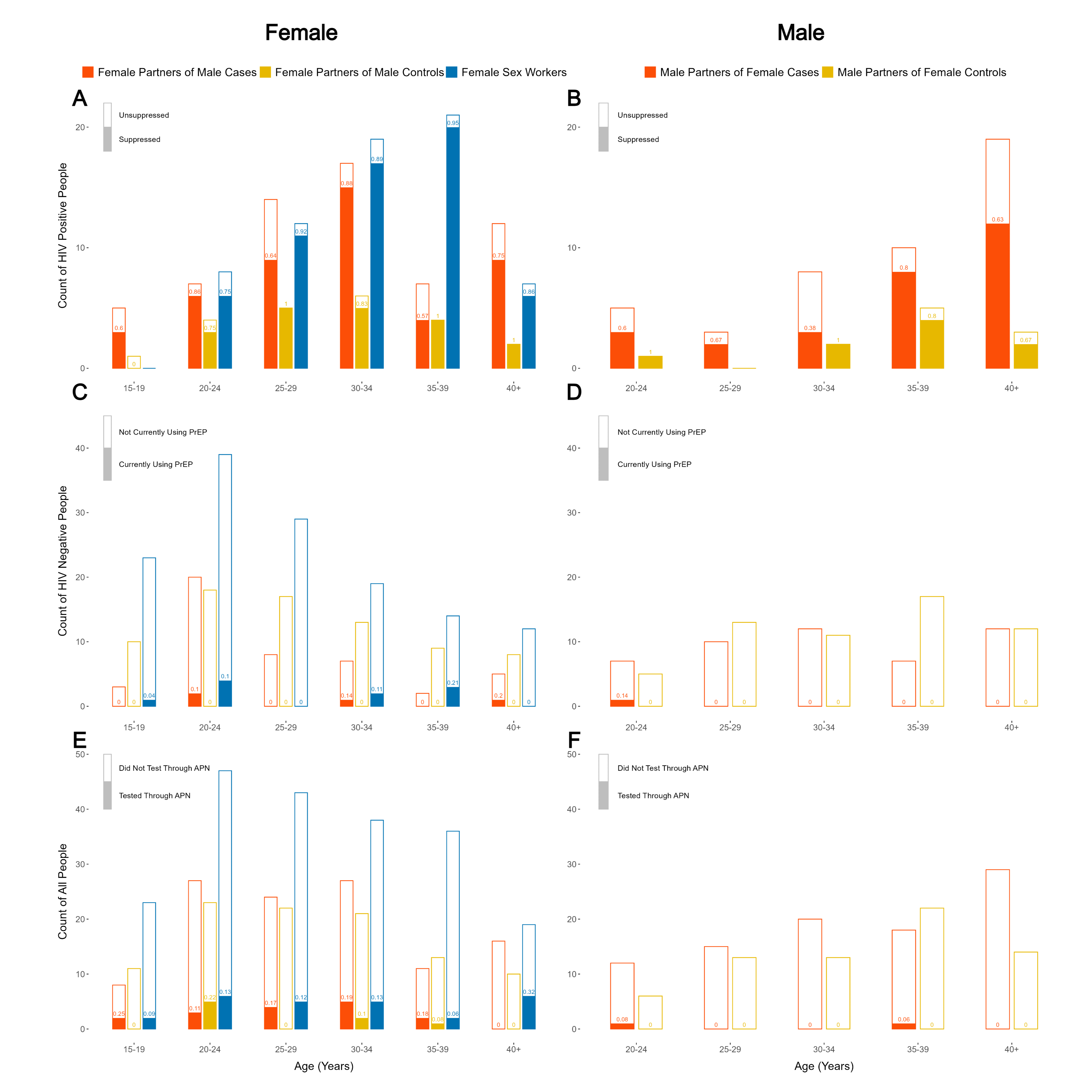

### Supplemental Figure 6: Distribution of HIV Viral Suppression, PrEP Use, and Assisted Partner Notification Testing Among Sexual Partners of Incident HIV Cases and Controls

*Panels A and B show the total number of HIV-positive partners among female partners of male participants and FSWs (A) and male partners of female participants (B). Light bars indicate the total number of HIV-positive partners, and dark bars indicate the subset who were virally suppressed (<200 copies/ml). Panels C and D show the number of HIV-seronegative partners; light bars indicate the total number of HIV-negative partners and dark bars indicate the subset currently using PrEP. Panels E and F show the total number of enrolled partners (light bars) and the subset who tested for HIV through assisted partner notification (APN) (dark bars). Female partners of male cases, female partners of male controls, and female sex workers (FSW) reported from male index participants are shown in panels A, C, and E. Male partners of female cases and controls are shown in panels B, D, and F. Results are stratified by partner age group.*

### Supplemental Table 4: Crude and Adjusted Matched Case-Control Odds Ratios from Conditional Logistic Regression, Overall and Stratified by Sex

| **Risk factor** | **Cases**  **N (%)** | **Controls**  **N (%)** | **Matched Odds Ratio (95% CI)** | **Adjusted* Odds Ratio (95% CI)** |
| --- | --- | --- | --- | --- |
| ***Demographics*** |  |  |  |  |
| Previously married | 53 (32.3%) | 24 (14.6%) | 3.07 (1.68, 5.61) | 3.00 (1.60, 5.62) |
| *Men* | *25 (31.6%)* | *11 (13.9%)* | *3.33 (1.34, 8.30)* | *3.37 (1.32, 8.58)* |
| *Women* | *28 (32.9%)* | *13 (15.3%)* | *2.87 (1.29, 6.43)* | *2.69 (1.15, 6.3)* |
| Education: secondary+ | 46 (28.0%) | 74 (45.1%) | 0.42 (0.25, 0.7) | 0.43 (0.25, 0.74) |
| *Men* | *22 (27.8%)* | *32 (40.5%)* | *0.52 (0.25, 1.09)* | *0.52 (0.24, 1.13)* |
| *Women* | *24 (28.2%)* | *42 (49.4%)* | *0.33 (0.16, 0.71)* | *0.35 (0.16, 0.78)* |
| ***Behavioral*** |  |  |  |  |
| Circumcision | 42 (54.5%) | 53 (68.8%) | 0.56 (0.29, 1.08) | - |
| *Men* | *43 (54.4%)* | *57 (67.1%)* | *0.56 (0.29, 1.08)* | *0.61 (0.30, 1.24)* |
| Consistent condom use | 77 (49.7%) | 97 (62.6%) | 0.57 (0.35, 0.91) | 0.73 (0.43, 1.23) |
| *Men* | *25 (31.6%)* | *40 (50.6%)* | *0.42 (0.20, 0.87)* | *0.49 (0.22, 1.06)* |
| *Women* | *52 (61.2%)* | *57 (67.1%)* | *0.73 (0.38, 1.38)* | *1.05 (0.49, 2.27)* |
| Age-disparate relationship (5 yrs) | 87 (58.8%) | 76 (51.4%) | 1.42 (0.84, 2.39) | 1.38 (0.79, 2.43) |
| *Men* | 47 (62.7%) | 39 (52.7%) | *1.90 (0.88, 4.09)* | *2.14 (0.92, 4.98)* |
| *Women* | 40 (54.8%) | 37 (50.0%) | *1.07 (0.52, 2.22)* | *0.92 (0.41, 2.05)* |
| Generational relationship (10 yrs) | 36 (24.3%) | 30 (20.5%) | 1.26 (0.69, 2.31) | 1.06 (0.56, 2.04) |
| *Men* | 22 (29.3%) | 15 (20.5%) | *2.00 (0.81, 4.96)* | *1.85 (0.7, 4.89)* |
| *Women* | 14 (19.2%) | 15 (20.5%) | *0.83 (0.36, 1.93)* | *0.64 (0.26, 1.61)* |
| Alcohol use before sex | 61 (48.0%) | 39 (30.7%) | 2.37 (1.32, 4.26) | 2.01 (1.07, 3.77) |
| *Men* | 34 (58.6%) | 26 (44.8%) | *2.00 (0.86, 4.67)* | *1.46 (0.57, 3.75)* |
| *Women* | 27 (39.1%) | 13 (18.8%) | *2.75 (1.22, 6.18)* | *2.47 (1.04, 5.86)* |
| Partner alcohol use before sex | 70 (55.1%) | 49 (38.6%) | 1.95 (1.17, 3.27) | 1.74 (1.00, 3.00) |
| *Men* | 28 (48.3%) | 22 (37.9%) | *1.46 (0.72, 2.96)* | *1.17 (0.54, 2.53)* |
| *Women* | 42 (60.9%) | 27 (39.1%) | *2.67 (1.24, 5.74)* | *2.54 (1.13, 5.71)* |
| ***Sexual Network*** |  |  |  |  |
| ≥1 enrolled partner with HIV | 93 (56.7%) | 28 (17.1%) | 11.83 (5.14, 27.23) | 11.81 (4.93, 28.28) |
| *Men* | *50 (63.3%)* | *18 (22.8%)* | *17.00 (4.08, 70.76)* | *16.16 (3.74, 69.87)* |
| *Women* | *43 (50.6%)* | *10 (12.1%)* | *9.25 (3.30, 25.95)* | *9.49 (3.15, 28.63)* |
| ≥1 enrolled partner with viremia | 32 (19.5%) | 5 (3.1%) | 7.75 (2.74, 21.96) | 10.11 (3.21, 31.81) |
| *Men* | *15 (18.4%)* | *3 (3.4%)* | *5.00 (1.45, 17.27)* | *6.97 (1.72, 28.31)* |
| *Women* | *17 (18.2%)* | *2 (2.0%)* | *16.00 (2.12, 120.65)* | *21.63 (2.44, 191.79)* |
| ≥2 reported total partners | 114 (69.5%) | 60 (36.6%) | 7.00 (3.48, 14.07) | 5.81 (2.83, 11.9) |
| *Men* | *68 (86.1%)* | *46 (58.2%)* | *5.40 (2.08, 14.02)* | *4.62 (1.75, 12.21)* |
| *Women* | *46 (54.1%)* | *14 (16.5%)* | *9.00 (3.20, 25.29)* | *7.73 (2.62, 22.84)* |
| ≥1 reported FSW partner | 34 (20.7%) | 5 (3.1%) | 15.50 (3.71, 64.77) | - |
| *Men* | *34 (43.0%)* | *5 (6.3%)* | *15.50 (3.71, 64.77)* | *14.57 (3.35, 63.3)* |
| ≥2 reported female non-sex-worker partners | 57 (34.8%) | 44 (26.8%) | 2.00 (1.03, 3.89) | - |
| *Men* | *57 (72.2%)* | *44 (55.7%)* | *2.00 (1.03, 3.89)* | *1.91 (0.95, 3.84)* |

*Adjusted by marital status (previously married or not) and education (secondary or higher). Network variables also adjusted by number of partners reported.

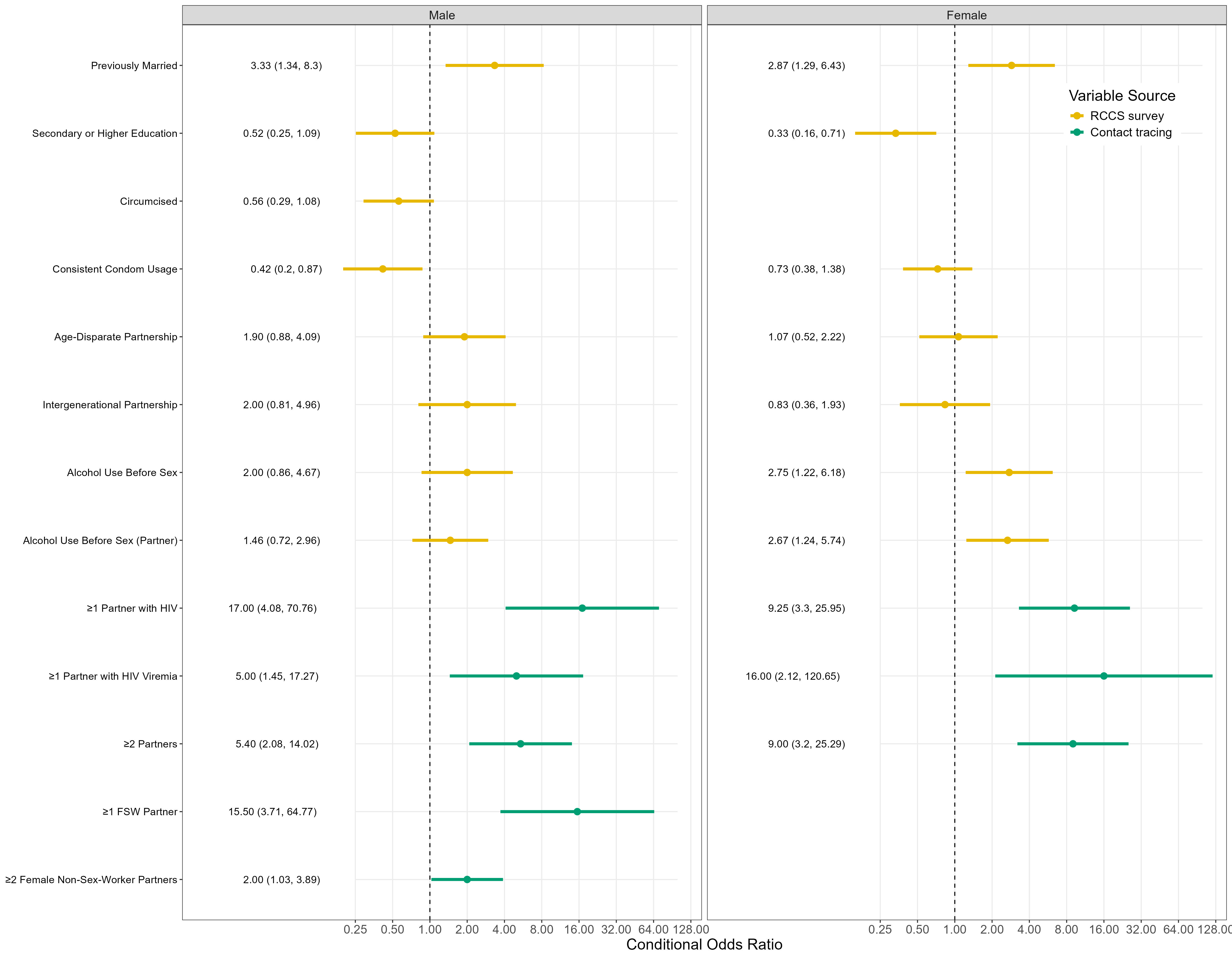

### Supplemental Figure 7: Demographic, Behavioral, and Network Predictors of Incident HIV, Stratified by Sex

*Points represent crude conditional odds ratio estimates, while lines represent the associated 95% confidence interval. Yellow points and intervals indicate variables that were collected during the Rakai Community Cohort Study (RCCS) survey or were constructed using those variables, while green points and intervals represent variables collected during contact tracing after enrollment in this case-control study. All variables were dichotomous, with the comparison group being the inverse of the exposed group. Previously married includes those widowed and divorced, and the comparison group includes people who are married and never married. Consistent condom usage includes those who always use condoms with casual partners or who have no casual partners. An age disparate partnership is defined as a partnership where the man was 5 or more years older than the woman and an intergenerational partnership was one where the man was 10 or more years older. Only enrolled partners could be tested for HIV and HIV viremia; however, total number of partners includes untraceable reported partners. Condom use and partner ages were obtained from partner block data from the Rakai Community Cohort Study, and collapsed into dichotomous variables as described in the Supplemental Methods.*

### Supplemental Table 5: Transmission Model Main and Sensitivity Analysis Results

| **Purpose** | **Transmission Model** | **Imputation Method for FSW Partners of Cases** | **Percentage of Infections from FSW**  **(95% CI)** | | |
| --- | --- | --- | --- | --- | --- |
|  |  |  | *All Cases* | *Male Cases* | *Male Clients* |
| Sensitivity | Network | Random from partners of same sex/arm, venue-enrolled FSW | 13.9  (12.9, 14.9) | 28.9  (26.7, 31.0) | 67.2  (62.0, 72.1) |
| Main result | Network | Weighted random from partners of same sex/arm, venue-enrolled FSW  (HIV prevalence = 64.7%*) | 16.6  (15.2, 17.7) | 34.4  (31.5, 36.8) | 80.0  (73.2, 85.4) |
| Sensitivity | Network | Weighted random from partners of same sex/arm, venue-enrolled FSW  (HIV prevalence = 80.1%^) | 18.0  (17.0, 18.9) | 37.4  (35.2, 39.2) | 86.9  (81.9, 91.1) |
| Sensitivity | Network | Weighted random from partners of same sex/arm, venue-enrolled FSW  (HIV prevalence = 46.3%^#^) | 13.8  (12.3, 15.3) | 28.7  (25.5, 31.7) | 66.7  (59.2, 73.6) |
| Sensitivity | Simple | Random from partners of same sex/arm, venue-enrolled FSW | 13.1  (11.0, 15.8) | 27.1  (22.9, 32.9) | 63.1  (53.1, 76.3) |
| Sensitivity | Simple | Weighted random from partners of same sex/arm, venue-enrolled FSW  (HIV prevalence = 64.7%*) | 14.5  (11.9, 17.8) | 30.1  (24.7, 37.0) | 70.0  (57.3, 85.9) |
| Sensitivity | Simple | Weighted random from partners of same sex/arm, venue-enrolled FSW  (HIV prevalence = 80.1%^) | 15.6  (13.0, 18.2) | 32.4  (27.0, 37.8) | 75.4  (62.8, 87.9) |
| Sensitivity | Simple | Weighted random from partners of same sex/arm, venue-enrolled FSW  (HIV prevalence = 46.3%^#^) | 12.9  (9.5, 15.6) | 26.8  (19.7, 32.4) | 62.2  (45.8, 75.3) |

** 64.7% is the estimated HIV prevalence among enrolled FSW partners of male cases*

*^ 80.1% is upper limit for the 80% confidence interval of HIV prevalence among enrolled FSW partners of male cases*

*^#^ 46.3% is the lower limit for the 80% confidence interval of HIV prevalence among enrolled FSW partners of male cases*

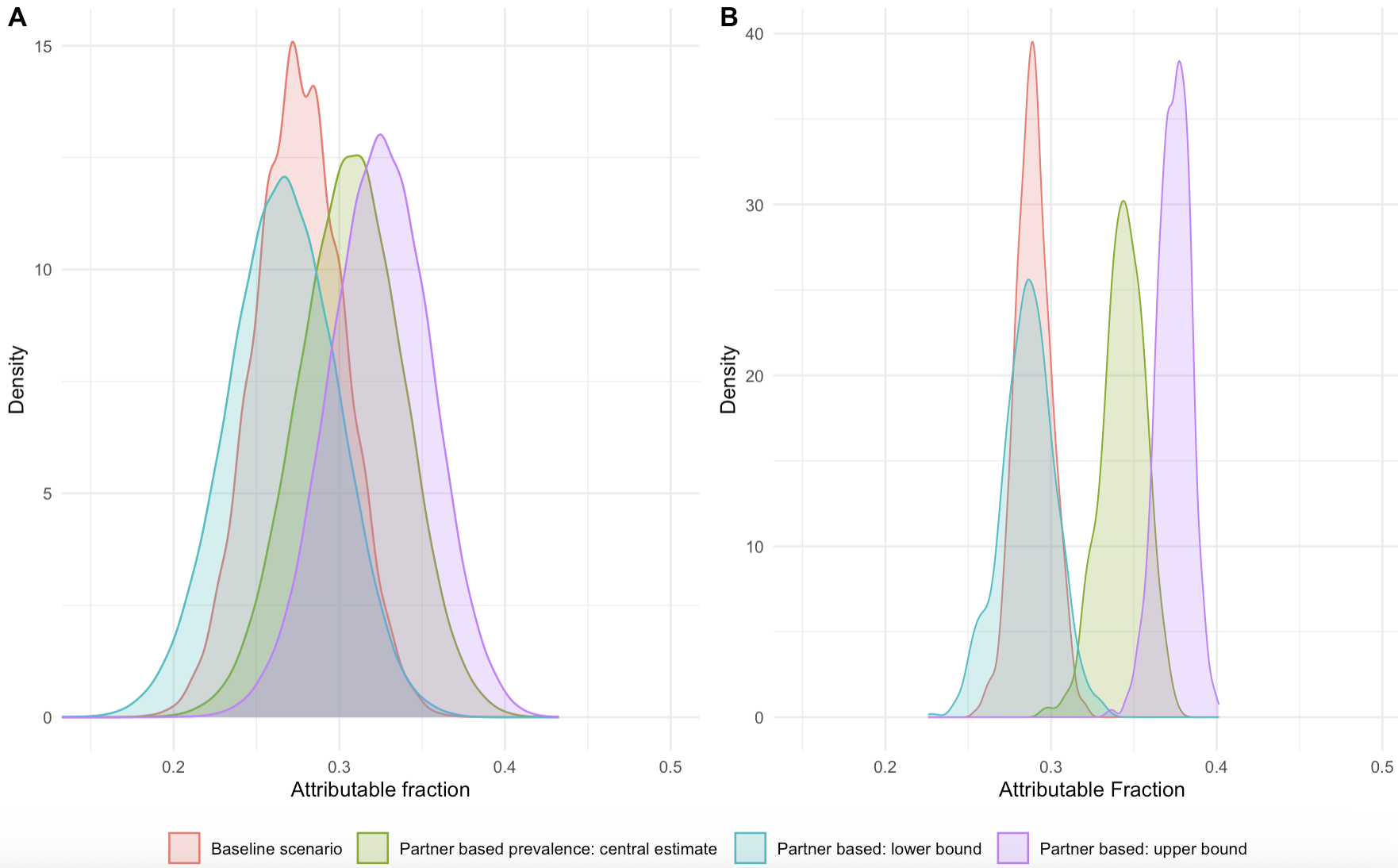

### Supplemental Figure 8: Transmission Model Main and Sensitivity Analysis Posterior Distributions

*Panels A and B show the distribution of transmission-model-derived estimates for the proportion of incident infections among men attributable to sexual partnerships with female sex workers, using different imputation methods, fully described in the Supplemental Methods, where the baseline scenario is imputation using partners of the same sex/arm and venue-enrolled FSW and other methods use a weighted random sampling to achieve an expected HIV prevalence among imputed partners. Panel A) displays the results from the simple transmission model while panel B) displays the results from the network transmission model.*
