## Supplementary Methodology for "HIV Transmission in a Declining African Epidemic"

**RCCS Survey Methods**

The RCCS is an ongoing population-based cohort established in 1994 that conducts household censuses and surveys in 34 agrarian, semi-urban trading and Lake Victoria fishing communities in in four districts (Kyotera, Masaka, Lyantonde, Rakai) in southern Uganda. It is administered by the Rakai Health Sciences Program (RHSP). Study community boundaries were established in 1994, and communities are surveyed sequentially each round. In each community, the RCCS first conducts a household census that enumerates all household members regardless of physical presence at the time of the enumeration. Individuals aged 15 years and older who reside in that community are eligible for participation, and a survey is administered to all eligible individuals who are present within two weeks of the census. The survey includes questions related to demographics, occupation, sexual behaviors, and health services utilization, including engagement with HIV programs. Participants also are asked to report on their four most recent sexual partners in the past year using an egocentric partner network module that collects partnership-level information on relationship type (marital, casual), age of partner, occupation, frequency of condom use (none, consistent, inconsistent use), and self and partner alcohol consumption.

**Regional HIV control Program**

During the study period, routine HIV prevention, testing, and treatment services in districts that include RCCS communities were delivered primarily through the U.S. President’s Emergency Plan for AIDS Relief (PEPFAR), implemented by the United States Centers for Disease Control and Prevention (US CDC) through regional implementing partners. The RCCS study communities fall within the Masaka regional HIV program, which covers 12 districts in south-central Uganda. From 2004 through 2023, the Rakai Health Sciences Program (RHSP) served as the primary PEPFAR implementing partner in this region, supporting HIV testing, antiretroviral therapy, and prevention services including pre-exposure prophylaxis (PrEP) for approximately 2.6 million people across 196 clinical sites.

As part of routine key population outreach activities, sex work venues were mapped approximately every two years to support targeted HIV testing, condom distribution, and PrEP delivery. These venue lists were maintained by implementing partners as part of routine program monitoring and outreach planning. In 2023, following completion of the RHSP cooperative agreement, responsibility for HIV service implementation in the region transitioned to the Infectious Diseases Institute (IDI), with services progressively being integrated into government health facilities under the oversight of the Ugandan Ministry of Health. In this study, venues identified during field activities were compared with previously mapped venues from the regional program.

**HIV Deep Sequencing and Genomic Linkage Analysis**

HIV deep sequencing was done on two groups of participants: 1) participants viremic at enrollment; and 2) participants who were suppressed at enrollment but had a blood sample with detectable virus collected during a previous RCCS visit. Deep sequencing was done on the Illumina platform using the veSEQ-HIV protocol (1). Consensus sequences were generated from deep sequences as previously described (1). Briefly, shiver (2) was used to process each sample as follows: sequencing reads were quality-trimmed and bioinformatically host-depleted before being de novo assembled, scaffolded against a curated set of HIV genomes from the Los Alamos HIV database, and re-mapped to the resulting sample-specific reference genome. Finally, consensus sequences were called at a minimum depth of 5 deduplicated reads as previously described (1).

We assessed genomic linkage between reported partners with HIV to evaluate whether epidemiologic links were supported by viral sequence data. Tamura-Nei (TN93) distances were calculated for the whole genome, and separately for *gag*, *pol*, and *env*, with linkage defined at region-specific thresholds (*pol* < 2%, *gag* < 3%, or *env* < 5%). If a pair lacked sufficient overlap in gene regions, an overall distance of 4.5% or less was considered evidence of linkage. Thresholds were selected based on region-specific mutation rates (3), similar thresholds in the literature (4–9), and examination of the distributions of genetic distances in our data. Usage of multiple region-specific and whole genome thresholds were intended to allow linkage to be robust to recombination and partial sequencing errors.

**Imputation and Transmission Modeling Methodology**

*Partner Imputation*

Not all partners of index participants were enrolled in the study. For the transmission model, we therefore imputed key characteristics of unenrolled partners, including age, number of sexual partners, and HIV serostatus. Occupation (female sex worker [FSW] vs. non-sex-worker [non-SW]) was available for all reported partners because sex work status was defined based on index participant report. Because imputation decisions could influence model results, we implemented four imputation strategies, detailed in the table below. Imputation strategies for non-SW partners of FSW partners of controls were consistent irrespective of strategy. The imputation strategy for FSW partners of cases changes: in strategy 1, the imputation is done randomly from venue-enrolled FSW while in strategies 2-4 imputation is done through weighted random sampling of venue-enrolled FSW to achieve a desired expected HIV prevalence. HIV prevalence targets include the main estimate and 80% confidence limits from the estimate of HIV prevalence among enrolled FSW partners of male cases: 64.7 (46.3, 80.1).

| **Strategy** | **Imputation of non-SW partners cases and controls** | **Imputation of FSW partners of controls** | **Imputation of FSW partners of cases** |
| --- | --- | --- | --- |
| 1 | Replacements randomly drawn from enrolled non-SW partners of indexes in the same arm and of the same sex | Replacements randomly drawn from venue-enrolled FSW and enrolled FSW partners of indexes in the same arm and of the same sex | |
| 2 | Replacements randomly drawn from enrolled non-SW partners of indexes in the same arm and of the same sex | Replacements randomly drawn from venue-enrolled FSW and enrolled FSW partners of indexes in the same arm and of the same sex | Replacements randomly drawn from venue-enrolled FSW and enrolled FSW partners of indexes in the same arm and of the same sex, but weighted to achieve an expected HIV prevalence of 64.7% |
| 3 | Replacements randomly drawn from enrolled non-SW partners of indexes in the same arm and of the same sex | Replacements randomly drawn from venue-enrolled FSW and enrolled FSW partners of indexes in the same arm and of the same sex | Replacements randomly drawn from venue-enrolled FSW and enrolled FSW partners of indexes in the same arm and of the same sex, but weighted to achieve an expected HIV prevalence of 80.1% |
| 4 | Replacements randomly drawn from enrolled non-SW partners of indexes in the same arm and of the same sex | Replacements randomly drawn from venue-enrolled FSW and enrolled FSW partners of indexes in the same arm and of the same sex | Replacements randomly drawn from venue-enrolled FSW and enrolled FSW partners of indexes in the same arm and of the same sex, but weighted to achieve an expected HIV prevalence of 46.3% |

*Transmission Modeling*

We developed two transmission models (simple and network-based) to estimate the proportion of incident cases arising from transmission from FSWs, henceforth referred to as the model-based attribution fraction (MAF). Inherent in both models is the assumption that the index did not transmit HIV to their partners, which may be reasonable given that these were recently infected cases, that the index acquired HIV from one of their reported partners, and that untraceable partners of index participants are different people. For simplicity in both models, we did not consider viral load of partners as a predictor of transmission.

The simple model assumed that all partners with HIV had an equal chance of infecting the index. Over 2000 simulations, an HIV-positive partner was randomly chosen as the transmitting partner for each index, and the proportion of these selected partners that were FSWs was calculated.

The network-based model is a Bayesian additive cumulative hazard acquisition model with dyad-specific transmission weights derived from partner covariates. Computing the MAF required the specification of the full likelihood for the HIV status of each partner of the index, attribution of the index’s cumulative transmission hazard to FSW and non-FSW dyads and then determining the proportion of the hazard attributable to dyads with a FSW partner. The assumptions of this model are the following:

- Each index and their local network are independent, and the full likelihood factorizes across indexes as a product of index-specific likelihood contributions.
- Partner HIV statuses are independent of the statuses of other partners.
- Each partner’s cumulative hazard of acquiring HIV was determined by (i) a social exposure component (i.e., number of partners over a year) and (ii) a per-partnership behavioral component.
- Only the behavioral component was assumed to affect the dyad-level transmission hazard from partner to index. This behavioral multiplier, together with a constant reflecting sex-specific biological differences and unmeasured index-level heterogeneity, determines the transmission hazard.

The covariates used in the model are partner HIV serostatus (positive and negative), age (<25 years and ≥25 years), number of sexual partners (<3 and ≥3), and occupation (FSW vs non-SW). The model included four parameters for partner HIV acquisition: baseline risk of acquisition of HIV, and acquisition multiplicative hazards for age, number of partners, and occupation. Each partner who identified as a FSW was assumed of have ≥3 sexual partners.

Each partner had a cumulative hazard of acquiring HIV which depends on a) a baseline population-level risk, b) the current age of the partner (reflecting duration of exposure), c) the number of partners of the partner (capturing exposure frequency), and d) partner intrinsic behavioral risk propensity. This last component is assumed to capture partner-level behavioral risk that operates across all of that partner’s sexual contacts. In other words, we assumed that behavioral factors influencing a partner’s probability of acquiring HIV (e.g., patterns of condom use or other high-risk practices) also proportionally influence the hazard of transmitting HIV within the dyad with the index. Because partnership-specific behavioral data were not available at the individual level, we treat this behavioral component as a partner-level multiplier that applies uniformly across that partner’s contacts, including the index.

For each index, the likelihood of the observed HIV status of their partners is the product of individual probabilities of acquiring HIV. For each negative partner p, this amounts to $e^{-\Lambda_{p}T_{p}}$ for that partner, with $\Lambda_{p}=\pi\beta_{N}^{p}\beta_{a}^{p}\beta_{o}^{p}$, where $\pi$ is the baseline hazard, $\beta_{N}^{p}$ is the relative hazard of acquisition due to the number of partners, $\beta_{a}^{p}$ is the relative hazard of acquisition due to age, $\beta_{o}^{p}$ is the behavioral hazard due to occupation, and $T_{p}$ is the time since first exposure to HIV in the lifetime of the partner. The complementary to 1 holds for each positive partner. We assume that $\pi\beta_{a}^{p}T_{p}\sim\tilde{\pi}\beta_{a}^{p}$ for all the partners. The likelihood of observing the sero-status of a configuration of partners in a sub-network around an index i can be written as

${L_{i}= \Pi}_{p} e^{-\Lambda_{p}T_{p}} \Pi_{n} (1-e^{-\Lambda_{n}T_{n}}$), where p stands for HIV positive partners and n for HIV negative ones.

The likelihood for all the sub-networks is just $L=\prod_{i} L_{i}$.

Finally, the relative hazard of HIV transmission originating from FSWs to each index i is $RH_{i}= \frac{\sum_{\boldsymbol{j\in FS}\boldsymbol{W}_{\boldsymbol{i}}} {\boldsymbol{\beta}^{\mathbf{j}}}_{\boldsymbol{o}}}{\sum_{\boldsymbol{j\in partners}_{\boldsymbol{i}}} {\boldsymbol{\beta}^{\mathbf{j}}}_{\boldsymbol{o}}}$, and the global MAF was calculated as $p_{sw}=\frac{1}{N}\begin{aligned} \sum^{N} \\ i=1 \end{aligned}RH_{i} ,$ where the sum goes over the N indexes.

Independent log-normal priors were placed on the multiplicative parameters (normal priors on the log-scale), and posterior inference was performed via MCMC using Stan. The full code for the implementation is available at https://github.com/FraDl89/Hard-to-reach-model.
